# Spillover Benefit of Pre-Exposure Prophylaxis for HIV Prevention: Evaluating the Importance of Effect Modification using an Agent-Based Model

**DOI:** 10.1101/2022.02.22.22271345

**Authors:** Ashley L. Buchanan, Carolyn J. Park, Sam Bessey, William C. Goedel, Eleanor J. Murray, Samuel R. Friedman, M. Elizabeth Halloran, Natallia V. Katenka, Brandon D.L. Marshall

## Abstract

We developed an agent-based model using a trial emulation approach to quantify effect measure modification of key variables on spillover effects of pre-exposure prophylaxis (PrEP) among men who have sex with men (MSM) in Atlanta, GA. PrEP may impact not only the individual prescribed, but also their partners and beyond, known as spillover. We simulated a two-stage randomized trial with eligible components (≥3 agents and ≥1 HIV+ agent) first randomized to intervention or control (no PrEP). Within intervention components, agents were randomized to PrEP with intervention coverage of 70%, providing insight into a high PrEP coverage strategy. We evaluated effect modification by component level characteristics and estimated spillover effects using an extension of randomization-based estimators. We observed an attenuation of the spillover effect when agents were in components with a higher prevalence of either drug use or bridging potential (if an agent acts as a mediator between ≥2 connected groups of agents). The estimated spillover effects were larger in magnitude among components with either higher HIV prevalence or greater density (number of existing compared to all possible partnerships). Consideration of effect modification is important when evaluating the spillover of PrEP among MSM.

## Introduction

Men who have sex with men (MSM) remain a population at high risk for HIV infection in the United States (US) and face barriers to optimal use of HIV prevention modalities, such as pre-exposure prophylaxis (PrEP), particularly in the Southern US (1). Many MSM are embedded in sexual risk networks such that the intervention may not only impact the individual prescribed PrEP, but also benefit their partners and beyond, which is known as *spillover or dissemination* (2, 3). The magnitude and extent to which PrEP confers spillover benefits to persons not prescribed themselves remains poorly understood.

Current PrEP prescribing guidelines from the Centers for Disease Control and Prevention (CDC) focus primarily on individual HIV risk behaviors, missing features of the sexual networks and larger context (4, 5). The package insert for brand-name PrEP medications does include assessment of HIV prevalence in individual’s social networks and other factors that increase vulnerability, including incarceration, exchanging sex for commodities, and drug use (6). However, the impact of features of the sexual network on spillover effects of PrEP remains largely unknown.

Causal inference using a potential outcomes framework (7) was proposed to estimate spillover effects in agent-based models (8), which are a type of individual-based microsimulation. Due to the complexity in the exposures resulting in spillover and various assumptions about the spillover mechanism, identification and estimation of causal effects is critical to ensuring the validity of the estimated spillover effects (9). Previous work employed an agent-based model to emulate a two-stage randomized trial to quantify the spillover effects of PrEP use among MSM in Atlanta, GA (10). That study demonstrated a spillover benefit of PrEP in the sexual networks of MSM among persons not assigned to PrEP themselves; however, this prior study did not consider effect modification. Possible effect modification of the spillover effects by component-level characteristics in a sexual network, such as HIV and drug use prevalence, which, if present, could be used to increase PrEP uptake and better allocate resources, such as increasing prescriber time with individuals whose treatment may benefit that individual and their partners (11). Components are defined as subsets of agents (individuals) connected through sexual partnerships in a sexual network, but not sharing partnerships with agents in other components.

Nationally, African American (AA) MSM have an estimated HIV prevalence (25%) more than double that of White (W) MSM (8%) (12). In Atlanta, GA, this difference was more pronounced with an estimated prevalence of 43% among AAMSM, compared to 13% among WMSM. Neither race-specific assortativity in sexual partnering nor sexual behaviors can fully explain these differences (13), highlighting that these disparities are driven by more distal structural forces (like poverty and incarceration), rather than individual behaviors. Despite these disparities in HIV prevalence, PrEP uptake among AAMSM remains limited (14, 15). Furthermore, several studies have demonstrated that the CDC criteria for prescribing PrEP poorly identified AAMSM who eventually seroconvert (5, 16). Evaluation of effect modification offers information on what types of sexual networks could benefit the most from an intervention with spillover benefit.

In this methodologically-focused study that performs causal inference with agent-based modeling, we evaluated effect modification of the spillover effect by component-level characteristics, including HIV and drug use prevalence and network features, specifically density (17) and bridging potential (18), in the context of a simulated two-stage randomized trial. Bridging potential is a measure of centrality of an individual agent where they could act as a mediator between two or more closely connected groups of agents. We adapted a previously calibrated model of PrEP uptake and HIV transmission among MSM in Atlanta to evaluate the spillover effects of PrEP on the outcome HIV incidence (10, 19, 20). We aimed to evaluate the magnitude and direction of possible spillover effects of PrEP use among MSM in Atlanta across predefined levels of the component-level effect modifiers to demonstrate the utility of causal inference methodology in agent-based modeling.

## Methods

### Model Setting and Simulated Trial

We used titan-model v2.1.0 (21) to simulate an agent-based model of PrEP uptake and HIV transmission among MSM in Atlanta that was calibrated using available published data (19, 20). We employed an iterative indirect approach following published guidelines (22). Outputs from the initial model setting were obtained and compared to surveillance data from the Georgia Department of Health (23) (see Supplementary Content Appendix 7). This agent-based model included 11,245 agents based on estimates that 5.4% of the adult male population in the wider Atlanta area were MSM (24). We then employed this calibrated model setting and demographics of this target population in our study; however, our aim was to simulate a randomized trial, rather than recreate the HIV epidemic in this setting. In this work, the simulated population was scaled to 17,440 agents to allow for enough components for the simulated randomized trial.

The agent-based model simulated a population of agents within a static sexual network; relationships and agents were initiated at population creation, and no agents entered or exited the model during the simulated trial for each run of the model. Agents were assigned demographics, sexual behavior characteristics, and HIV prevalence and treatment at model initiation, and components in the network were identified based on sexual partnerships formed at model initialization. Partner selection was a function of race and drug use class (25), which resulted in the generation of assortative sexual networks. This model used a ‘bottom-up’ approach for generating networks, where agents were assigned a number of partners, types of partners, then partnerships were selected through an iterative process (21). At model initialization for each model run, agents were assigned a target number of sexual partners. The total number of sexual partners per year was assumed to follow a negative binomial distribution with mean = 5 for AAMSM and mean = 7 for WMSM (13). The number of sexual acts per month within a partnership was assumed to follow a Poisson distribution, and each agent was assigned a total number of sexual acts per interval, based on a distribution of monthly number of sexual acts (12). For each agent, a pool of potential partners was created from all other agents seeking partners, and subsequently narrowed by sexual position and the agent’s assorting probabilities. Then, the agent selected partners from this generated pool to achieve its target number of partners.

The probability of HIV transmission for a sexual partnership depended upon the following factors: condom use; type of anal intercourse; PrEP use and adherence (if HIV-negative and assigned); knowledge of HIV seroconversion status, use of antiretroviral therapy (ART) and viral suppression (if HIV-positive). For partnerships with condomless sex, there were non-zero per-act probabilities of HIV transmission (per-act probability for condomless receptive anal intercourse was 1.38% and condomless insertive was 0.11%) (26). In addition, HIV-positive agents’ knowledge of their HIV status, ART utilization, and adherence to ART at model initiation modified the per-act probability of HIV transmission. At each monthly time interval, information on each agent and their component were recorded, including HIV status, HIV treatment and viral load, and number of sexual acts.

We simulated a two-stage randomized design among MSM in Atlanta evaluating the spillover effect of PrEP on the outcome cumulative HIV incidence by two years after randomization (10, 19, 20). Because we were interested in component-level effect modification of spillover effects to preserve interference sets, we required the components in the simulated trial to have at least three agents (one HIV+ agent) and individual agents had to be HIV-negative to receive PrEP. This inclusion criteria ensured that the components in the trial included individuals at risk for HIV infection and the network structure of the component was complex enough to measure centrality.

In the simulated two-stage randomized design, enrolled components were first randomized 1:1 to either a PrEP allocation strategy (“intervention” components) or no PrEP allocation (control components). Then, eligible agents in each “intervention” component were randomized to PrEP according to a specific coverage level defined by the assigned allocation strategy. We considered the scenario of 70% PrEP coverage in intervention components to provide insight into strategies with high PrEP coverage.

We assumed the same set of parameters for this study as for the previously published work (19). However, some key adaptations were made for this study (10). We assumed static sexual networks as defined at baseline were fixed over time in each simulated trial. The full sexual sociometric network was a set of smaller components where each agent belonged to only one component, and there were no partnerships between agents in different components. We also assumed no drop out (i.e., 100% retention in PrEP over the two years). These assumptions were necessary because the existing methods for evaluating spillover do not allow for time-varying components in the sexual network.

We added a “drug use” agent class, which was defined at model initialization and remained stable for the duration of the simulated trial. The prevalence of drug use was defined based on a review of relevant literature (27). Drug use was defined as self-reported use of cannabis, cocaine, amphetamines, methamphetamines, inhalant nitrites, heroin/ opioids, or benzodiazepines in the past 12 months and influenced PrEP adherence, condomless sex, and assortativity. Specifically, agents who were defined as using substances had a 35% lower probability of adherence to PrEP (25) and 20% higher probability of condomless sex (28). We assumed that 20% of substance-using agents mixed with other substance using agents.

Python software, version 2.7.12, along with the NumPy and NetworkX packages, was used for coding, testing, and performing sensitivity analyses of this model. The analysis of model output for this paper was generated using SAS software (version 9.4, Cary, NC, USA). R software, version 3.5.1, along with ggplot2, was used to produce figures. Additional information regarding parameter values, key model assumptions, data sources, and additional references are included in the Supplementary Material Appendices 1-8 (Table S1).

### Causal Inference Methods for Spillover

Let *M* be a baseline (pre-randomization) binary component-level variable (1 = presence of a component-level factor, 0 = absence of that factor). Let *Y* denote the agent-level outcome ascertained two years after randomization in the simulated trial. Given the two-stage randomized design, we expect exchangeability to hold at both the component and agent levels. We assume exchangeability within levels of M; that is, *Y*^*a*^ ⊥*A*|*M*. This means that the intervention components are comparable to control components, and within intervention components, agents randomized to the intervention are comparable to those randomized to the control, in expectation conditional on *M* (29).

Although static during follow-up, the component sizes at model initialization varied in each simulated trial due to the partnering algorithm, so we extended estimators (30) to evaluate effect modification by component characteristics. We assume *partial interference;* that is, an agent’s outcome is influenced only by others in the same component, but no agents outside the component. We also assume *stratified interference*, in which an agent’s potential outcome is dependent only on their own intervention assignment and the proportion of agents randomized to the intervention in their component (2). We make the usual additional assumptions required for causal inference (i.e., exchangeability, consistency, and positivity) (29). We assumed a Bernoulli allocation strategy for intervention assignment within each component (2).

To evaluate effect modification by network features, we first averaged agent-level characteristics for each component to determine the distribution across all components, then used this distribution to define a binary variable for each component. Let the indicator function *I* (*M*_*i*_ = *m*) = 1 if *M*_*i*_ = 1 and *I* (*M*_*i*_ = *m*) = 0, otherwise. Let *I*_*m*_ denote the number of components with *M*_*i*_ = *m* and 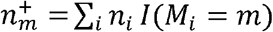 denote the number of individuals in stratum *M*_*i*_ = *m*. Among components with *M*_*i*_ = *m*, the spillover (i.e., disseminated or indirect) effect is

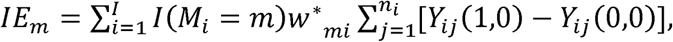

where 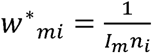 corresponds to component-weighted estimands and 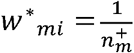 corresponds to individual-weighted estimands.

For the estimator, we computed the inverse probability weights conditional on M, then we estimated the causal effect within each level of the component-level variable. We employed both stabilized and unstabilized estimators in the analysis (30). These estimators are unbiased in a two-stage randomized design with a single allocation strategy and a control group (no agents were assigned to PrEP) (30). The estimators of the risk ratio of the spillover effect are defined analogously (30). Please see Supplementary Material Appendix 11 for additional details on the causal inference methods for evaluation of effect modification of spillover.

### Network Structure of Components

The network features considered were the bridging potential and density. The *network size* is defined as the number of nodes (i.e., agents) in the sexual network (17). An undirected *graph G* = (*V, E*) is a mathematical structure consisting of a set *V* of vertices or nodes (agents in our setting) and a set *E* of *edges* or *links* (i.e., sexual partnerships), where elements of *E* are unordered pairs {*u, v*} of distinct vertices *u, v* ∈ *V*.

A component’s density is defined as the proportion of observed connections in a component among the maximum number of possible connections in a component of the same size (17). The density *ρ*_*i*_ is defined as the fraction of those edges that are actually present for component *i*:

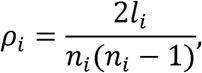

where *n*_*i*_ is the number of agents in the network component and *l*_*i*_ is the number of edges (or connections) in the component. A sexual network component that is both large and dense is more likely to have agents who engage in sexual partnerships within the network component. This is particularly problematic when a pair of agents are HIV serodiscordant.

Bridging potential (also known as effective size) measures the redundancy in an individual agent’s partnerships by examining the connections between their partners, providing a measure of centrality of an individual agent where they could act as a mediator between two or more closely connected groups of agents (18). For our study with unweighted and undirected graphs with one type of node, a simplified formula to compute bridging potential for agent *u*_*i j*_:

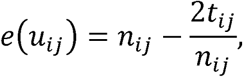

where *t*_*ij*_ is the number of edges (not including ties to agent *u*_*ij*_) and *n*_*ij*_ = *n*_*i*_ − 1 is the number of agents (excluding *u*_*ij*_) in component *i*. Bridging potential can be used to identify critical agents for interrupting HIV transmission chains in a network component. Agents with high bridging potential can act as gatekeepers in the network (17) and, in the context of HIV, are agents who divide relatively isolated groups of other agents. If these agents remain uninfected, for example, by adhering to a PrEP regimen, they would limit or slow the spread of infection in the population (Figure 1). Intervening on the HIV-negative agent with PrEP with no bridging potential would only protect that agent against HIV acquisition; however, intervening on the HIV-negative agent with PrEP with high bridging potential would protect that agent and the other HIV-negative agents in the component (31).

**Figure 1.**
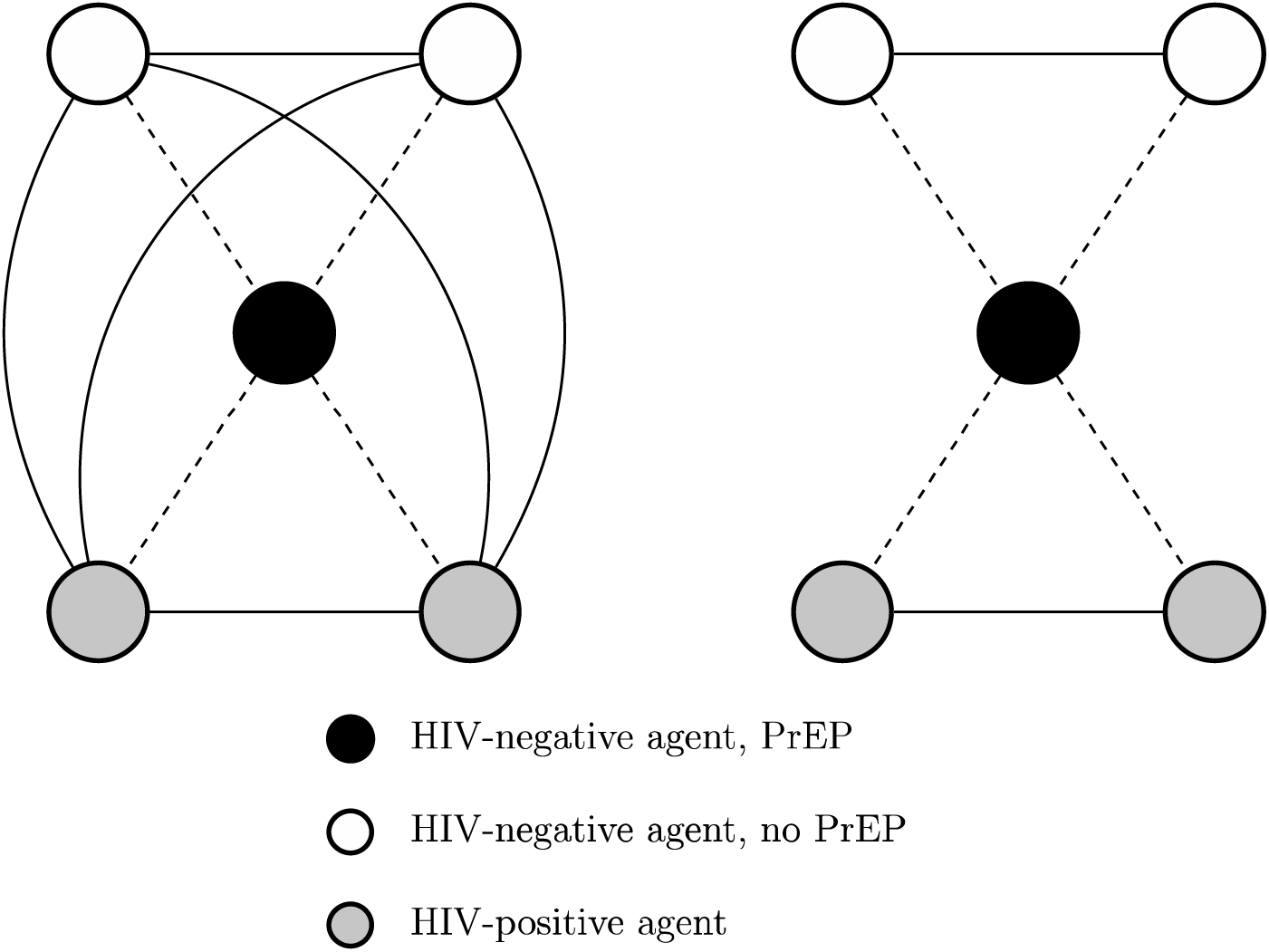
HIV-negative agent on PrEP who has no bridging potential (left) versus high bridging potential (right)^1^. ^1^Dashed lines represent sexual partnerships present in both components, while solid lines represent sexual partnerships in the fully connected component only (left). Adapted from (31).

### Outcome Measures

The primary outcome measure was cumulative HIV incidence over 24 months after randomization in the simulated trial, as measured by the number of incident HIV infections among those HIV-negative at the start of the trial (and reported as a proportion). We examined effect modification on both the ratio and difference scale using stabilized component-weighted estimators (30). We estimated the *spillover* effect within levels of the following component-level effect modifiers aggregated to the component level: HIV prevalence and drug use prevalence; and network characteristics: average density and average bridging potential. For each of the four effect modifiers, we defined binary variables based on the distribution of each variable (e.g.,≤ median vs. > median). These parameters were computed using nonparametric estimators for this setting, averaging across 1,000 simulations, along with 95% simulation intervals (SIs) (i.e., middle 95% of simulated output) to assess stochastic uncertainty (32).

### Sensitivity Analyses

The model results may depend on PrEP adherence and discontinuation among the agents randomized to PrEP. We performed one-way sensitivity analyses to evaluate the impact of modifying adherence to PrEP and discontinuation of PrEP. We quantified the effect modification, as specified above, across component-level factors. Specifically, we modified the proportion who were optimally adherent to PrEP in a monthly interval (i.e., 4 or more doses per week) to be 80% among WMSM and 50% among AAMSM (33). We considered a scenario where 10% of agents discontinued PrEP in each monthly interval during the two-year follow-up (34). We also conducted an additional sensitivity analysis in which PrEP coverage in the intervention components was set to 30% (on average), reflecting a lower coverage PrEP strategy.

## Results

In the two-stage simulated trial, there were an average of 3,947 agents per simulation and about 800 components per trial with an average component size of 5 agents (standard deviation (SD) = 3). Characteristics of components were balanced between the intervention and control components (Table 1). On average, HIV prevalence was approximately 34% in each component at enrollment (95% simulation interval (SI) = 33%, 36%). About 45% of agents were AA (95% SI = 42%, 47%) and prevalence of drug use was 35% in the components (95% SI = 34%, 37%), on average. Average bridging potential was 1.51 (SD = 0.17); therefore, an agent in the network component typically has either a single connection or lies between two other agents. Average density was 0.49 (SD = 0.17), implying about half of the possible connections were made in the simulated network.

**Table 1.** Characteristics of components at the time of enrollment into the simulated two-stage randomized trial with 70% PrEP coverage in the intervention group in an agent-based model representing men who have sex with men in Atlanta, Georgia, 2015-2017

| Characteristics | Summary Measure | Component Assignment |  |
| --- | --- | --- | --- |
|  |  | Intervention (n=1977.0) | Control (n=1969.5) |
| Number of Components |  | 401.1 | 399.7 |
| Average Component Size | Mean (SD) | 4.9 (3.2) | 4.9 (3.1) |
|  | Median (IQR) | 4 (2) | 4 (2) |
| HIV Prevalence |  | 34.2% | 34.3% |
| African American Race |  | 44.7% | 44.8% |
| Any substance use |  | 35.4% | 35.4% |
| Bridging potential | Mean (SD) | 1.5 (0.2) | 1.5 (0.2) |
|  | Median (IQR) | 1.5 (0.3) | 1.5 (0.3) |
| Density | Mean (SD) | 0.5 (0.2) | 0.5 (0.2) |
|  | Median (IQR) | 0.5 (0.3) | 0.5 (0.3) |

Note: results above are from 1,000 iterations of the agent-based model.

We evaluated the cumulative HIV incidence by 24 months of follow-up overall in the simulated trial and by four effect modifiers at the component level (Table 2). Overall, cumulative incidence among those in control components was 10% (95% SI = 8%, 12%), compared to 9% among agents randomized to no PrEP in the intervention components (95% SI = 6%, 12%) and 1% among agents randomized to PrEP in intervention components (95% SI = 0.5%, 2%). Among components with HIV prevalence above the median, the HIV cumulative incidence was about twice as high for all three groups compared to components with prevalence below the median. A similar but more modest trend was observed for density. For the effect modifiers drug use and bridging potential, the estimated cumulative incidence was comparable across levels of the effect modifiers.

**Table 2.** Cumulative incidence of HIV over two years of follow-up after two-stage randomization stratified by four modifiers (≤median vs. >median) among HIV-negative agents within PrEP intervention (70% coverage) and control components with 95% simulation intervals (SI) in an agent-based model representing among men who have sex with men Atlanta, Georgia,

| Effect Modifiers | Intervention Components |  |  |  |  |  | Control Components |  |  |
| --- | --- | --- | --- | --- | --- | --- | --- | --- | --- |
|  | Agents on PrEP |  |  | Agents Not on PrEP |  |  | Agents Not on PrEP |  |  |
|  | Total<br>Agents | HIV+<br>Agents | Cumulative<br>Incidence<sup>a</sup> | Total<br>Agents | HIV+<br>Agents | Cumulative<br>Incidence<sup>a</sup> | Total<br>Agents | HIV+<br>Agents | Cumulative<br>Incidence<sup>a</sup> |
| Overall | 909.8 | 10.4 | 0.01 | 389.7 | 35.4 | 0.09 | 1295.7 | 125.2 | 0.10 |
| Substance Use |  |  |  |  |  |  |  |  |  |
| Among $M = 0$ | 490.6 | 6.4 | 0.01 | 210.2 | 21.1 | 0.10 | 698.9 | 74.3 | 0.11 |
| Among $M = 1$ | 419.2 | 4.0 | 0.01 | 179.5 | 14.4 | 0.08 | 596.8 | 51.0 | 0.09 |
| HIV Prevalence |  |  |  |  |  |  |  |  |  |
| Among $M = 0$ | 700.1 | 6.0 | 0.01 | 300.1 | 21.6 | 0.07 | 998.0 | 77.5 | 0.08 |
| Among $M = 1$ | 209.7 | 4.4 | 0.02 | 89.7 | 13.8 | 0.16 | 297.7 | 47.7 | 0.16 |
| Bridging potential |  |  |  |  |  |  |  |  |  |
| Among $M = 0$ | 352.4 | 4.9 | 0.01 | 150.3 | 16.4 | 0.11 | 501.0 | 57.3 | 0.11 |
| Among $M = 1$ | 557.4 | 5.5 | 0.01 | 239.4 | 19.0 | 0.08 | 794.7 | 67.9 | 0.09 |
| Density |  |  |  |  |  |  |  |  |  |
| Among $M = 0$ | 719.3 | 7.5 | 0.01 | 308.1 | 25.8 | 0.08 | 1024.9 | 92.2 | 0.09 |
| Among $M = 1$ | 190.5 | 2.9 | 0.02 | 81.5 | 9.6 | 0.12 | 270.9 | 33.0 | 0.12 |

<sup>a</sup>Cumulative incidence presented as a proportion (number of HIV+ /total number).

Table 3 displays the estimated spillover effects of PrEP on cumulative incidence of HIV by 24 months on the ratio and difference scale stratified by the four binary modifiers. For the effect modifier drug use, the estimated spillover effects were similar among those components with a lower prevalence of drug use compared to higher drug use. For bridging potential, the estimated spillover effects were larger in magnitude on both the difference and ratio scales among components with lower average bridging potential, compared to those with higher bridging potential.

**Table 3.** Spillover effects of PrEP on cumulative incidence of HIV over two years of follow-up after two-stage randomization stratified by four effect modifiers (≤median vs. >median) among HIV-negative agents within PrEP intervention and control components with 95% simulation intervals (SI) in an agent-based model representing men who have sex with men, Atlanta, Georgia, 2015-2017 (n = 3,947)

|  | RD (95% SI) | RR (95% SI) |
| --- | --- | --- |
| **M=0** |  |  |
| Substance Use | -0.08 (-0.14, -0.03) | 0.47 (0.24, 0.78) |
| HIV Prevalence | -0.05 (-0.10, -0.01) | 0.56 (0.26, 0.91) |
| Bridging potential | -0.08 (-0.12, -0.03) | 0.44 (0.23, 0.72) |
| Density | -0.06 (-0.10, -0.01) | 0.58 (0.30, 0.94) |
| **M=1** |  |  |
| Substance Use | -0.07 (-0.14, 0.001) | 0.53 (0.20, 1.01) |
| HIV Prevalence | -0.13 (-0.22, -0.04) | 0.41 (0.18, 0.76) |
| Bridging potential | -0.04 (-0.03, 0.02) | 0.68 (0.31, 1.21) |
| Density | -0.08 (-0.14, -0.02) | 0.42 (0.17, 0.79) |

<sup>1</sup> Estimates calculated using component-weighted stabilized estimators (30) RD = Risk Difference; RR = Risk Ratio

The estimated spillover effects were larger in magnitude on both the difference and ratio scales among components with higher HIV prevalence compared to lower. Similar patterns were observed for network density. Figures 2 and 3 display the estimated spillover effects across 1,000 simulated trials on the difference and ratio scale. Consistent with the results in Table 3, the spillover effects among components with HIV prevalence above the median were larger in magnitude on both the difference and ratio scale, as compared to components with coverage below the median; however, these estimates had more variation across simulations for the difference scale.

**Figure 2.**
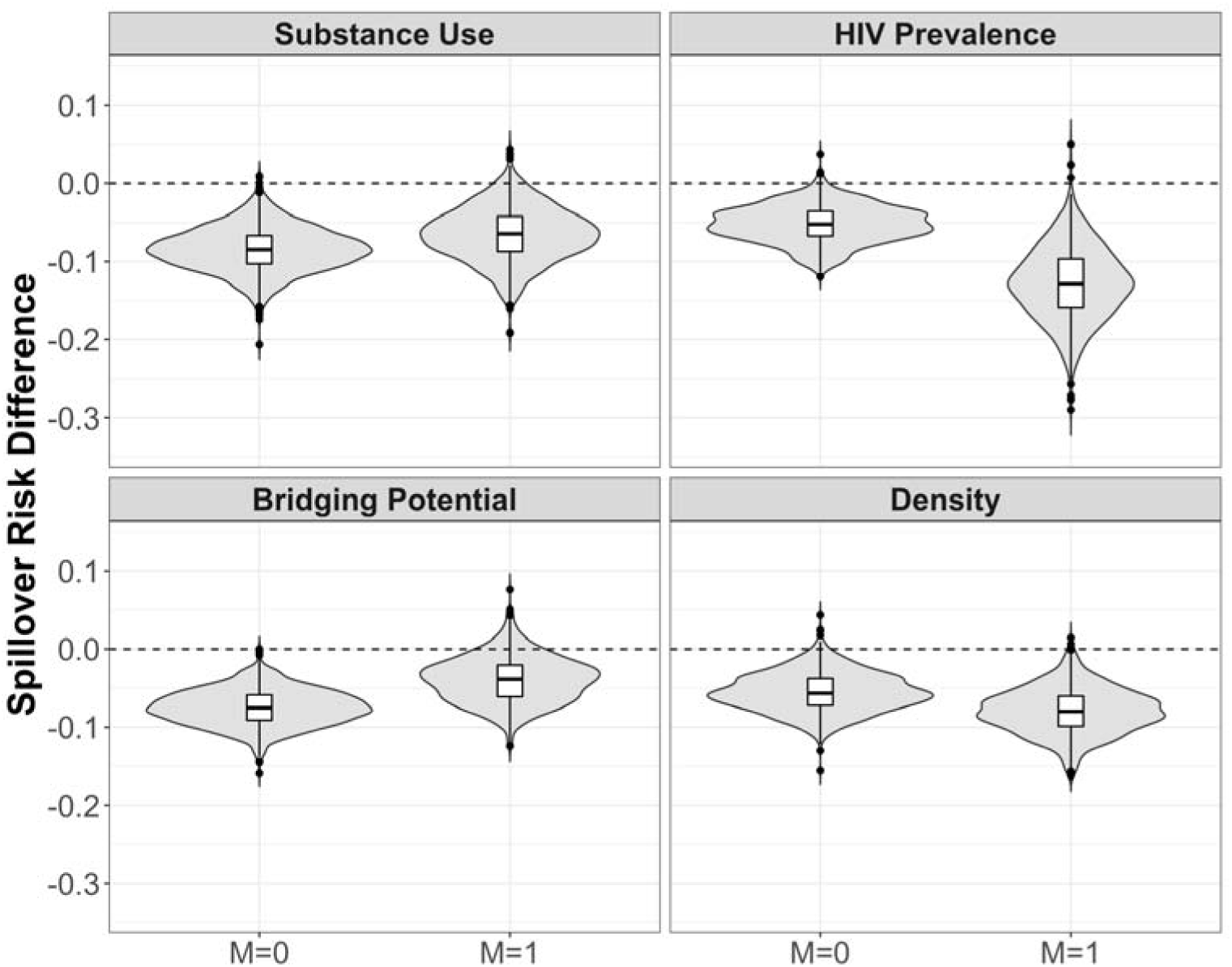
Estimated spillover risk difference of PrEP on cumulative incidence of HIV by effect modifiers^1,2^. ^1^M = 1 if prevalence above median (vs. M = 0 at or below median) among HIV-negative agents within PrEP intervention (70% coverage) and control components in two-stage randomized designs of a pre-exposure prophylaxis (PrEP) intervention with 70% coverage in an agent-based model representing men who have sex with men, Atlanta, Georgia, 2015-2017. ^2^ Lines within boxes, median values; box borders, interquartile ranges (75th and 25th percentiles); bars, 90th and 10th percentiles; points, outliers. Shaded shape represented the distribution of estimates and dashed lines represent the null value. (n = 3,947)

**Figure 3.**
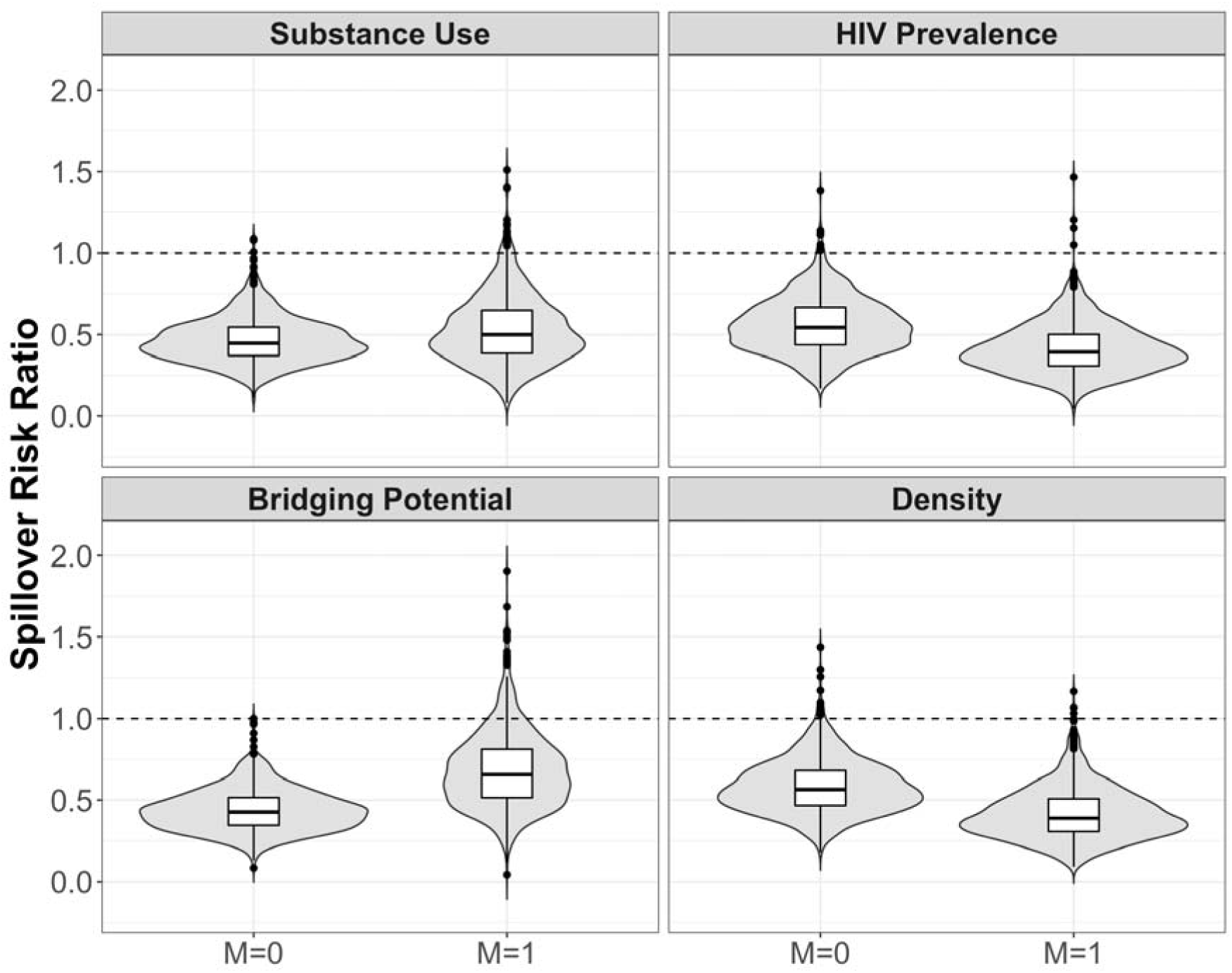
Estimated spillover risk ratio of PrEP on cumulative incidence of HIV by effect modifiers^1,2^. ^1^ M = 1 if prevalence above median vs. M = 0 at or below median) among HIV-negative agents within PrEP intervention (70% coverage) and control components in two-stage randomized designs of a PrEP intervention with 70% coverage in an agent-based model representing men who have sex with men, Atlanta, Georgia, 2015-2017. ^2^ Lines within boxes, median values; box borders, interquartile ranges (75th and 25th percentiles); bars, 90th and 10th percentiles; points, outliers. Shaded shape represented the distribution of estimates and dashed lines represent the null value. (n = 3,947)

For the sensitivity analysis, we simulated the trial with a 30% coverage of PrEP in the intervention components. In general, the estimated spillover effects were larger in magnitude with 70% intervention coverage, as compared to 30% coverage (Supplementary Material Appendix 9, Supplementary Material Tables S2-S6). We performed one-way sensitivity analyses to assess the impact of our model parameterization for PrEP adherence and PrEP discontinuation on model results for HIV cumulative incidence, focusing on two-stage randomized trials with 70% coverage allocation strategies in the intervention components. Interestingly, the estimated spillover effects were fairly robust to these one-way sensitivity analyses for either PrEP adherence or PrEP discontinuation and the results from both sensitivity analyses were comparable to the main analysis (Supplementary Material Appendix 10, Supplementary Material Tables S7-S18, Supplementary Material Figures S1-S4).

## Discussion

To leverage causal inference in agent-based models, we simulated a two-stage randomized trial to evaluate effect modification of spillover effects of PrEP among MSM in Atlanta, Georgia (19, 21). We found that the spillover effects were slightly stronger among components with a lower prevalence of drug use and lower average bridging potential. Interestingly, the spillover effects were stronger in components with *higher* HIV prevalence and also greater network density. In components with HIV prevalence above the median, the estimated spillover effect was a 59% reduction in HIV risk, as compared to only a 44% reduction in HIV risk among components with HIV prevalence at or below the median.

When there are more HIV-infected agents in a component, HIV-uninfected individuals have an increased risk of HIV acquisition, and thus benefit more by having partners who are on PrEP due to the reduction in HIV transmission risk from their partners’ concurrent partners (35). In fact, a component with no HIV-infected individuals at enrollment and no sexual risk connections outside the component has zero HIV risk, regardless of PrEP status, so these were excluded from our study. Furthermore, the estimated effects were stronger in denser components. When a component has more connections between the agents, there are more opportunities for PrEP to make a difference in terms of preventing HIV seroconversion of agents (36). In this model, drug use decreased PrEP adherence and condom use, as well as influenced the partnering algorithm. A lower prevalence of drug use likely resulted in less sexual risk behavior in a component possibly bolstering the spillover effect.

Particularly in the Southern US, uptake of PrEP services remains concerningly low among AAMSM (15). Recent efforts in Atlanta, GA, to expand access to PrEP through the county health department are notable; however, initiating and adhering to PrEP remain a significant challenge for successful delivery among AAMSM (34). Disparities in PrEP uptake may weaken the population-level impact on HIV incidence (20, 37). Many evaluations of the efficacy and effectiveness of PrEP focus on individual effect without consideration of the sexual risk network in which these individuals are embedded. With a better understanding of spillover effects and important effect modifiers, the delivery of PrEP interventions could be tailored to the most at-risk components, possibly mitigating disparities in PrEP uptake between WMSM and AAMSM. For example, network-based interventions could involve persons living with HIV referring their HIV-negative partners to PrEP. In fact, models such as the one employed here could be used to conduct preliminary evaluations of network-based PrEP interventions to inform subsequent delivery in the population (38, 39).

This simulated trial approach has several limitations. Motivated by the question of effect modification of spillover effects, we designed this study to include components both of meaningful size and at-risk for HIV. To employ existing causal inference methods, we assumed that the sexual network was static over time. This does not reflect the true underlying sexual networks among MSM in Atlanta, GA. In future work, we plan to develop an approach that allows for assessment of spillover with networks updated over time. We considered the entire sexual component to be an interference (i.e., spillover) set, which means that the intervention status of one agent in the component could possibly affect the outcomes of all other agents in the component. If a component is large or not well connected through sexual behaviors, this assumption may be dubious. A more realistic interference set might include an agent’s sexual partners and their partner’s partners. Future work should include an evaluation of different interference sets, for example, by considering an agent’s partners, known as nearest neighbors (40).

Careful consideration of effect modification of intervention effects with possible spillover is useful to inform the development of interventions that leverage network features. Persons not on PrEP may benefit from being in a network with higher PrEP coverage levels, and this benefit may be larger when component-level risk factors are more prevalent. Guidelines could encourage providers to ask about the PrEP status of sexual partners and encourage individuals on PrEP to recommend to others in their sexual network.

## Data Availability

Model code used to generate this data can be found at https://pph-collective.github.io/TITAN/. Complete reference for the TITAN model is S. Bessey, Mary McGrath, & Maximilian King. (2020, November 10). marshall-lab/TITAN: v1.2.4 (Version v1.2.4). Zenodo. http://doi.org/10.5281/zenodo.4266540. Data sets and post-modeling analysis code will be made available on Brown's Digital Repository.

## Availability of data and materials

Model code used to generate this data can be found at https://pph-collective.github.io/TITAN/. Complete reference for the TITAN model is S. Bessey, Mary McGrath, & Maximilian King. (2020, November 10). marshall-lab/TITAN: v1.2.4 (Version v1.2.4).

Zenodo. http://doi.org/10.5281/zenodo.4266540. Data sets and post-modeling analysis code will be made available on Brown’s Digital Repository.

## Authors’ Contributions

ALB conducted a literature review, developed the methodology, performed the statistical analysis, and contributed to manuscript writing. SB and CJP developed and performed the agent-based and statistical modeling and contributed to manuscript writing. WCG provided substantive guidance on the methodology development and contributed to the literature review and manuscript writing. EJM, MEH, and NVK contributed to the literature review, development of methodology and manuscript writing. SRF provided substantive guidance on the methodology development and contributed to the literature review and manuscript writing. BDLM provided guidance on the development and implementation of the agent-based models in this setting and contributed to the literature review and manuscript writing.

## Acknowledgement

Not applicable.

## Abbreviations

(ART): Antiretroviral therapy
(AA): African American
(CDC): Centers for Disease Control and Prevention
(HIV): Human immunodeficiency virus
(MSM): Men who have sex with men
(PrEP): Pre-exposure prophylaxis
(SI): Simulation interval
(SD): Standard deviation
(US): United States
(W): White

