## Supplemental File for "Spillover Benefit of Pre-Exposure Prophylaxis for HIV Prevention: Evaluating the Importance of Effect Modification using an Agent-Based Model"

**SUPPLEMENTARY MATERIAL**

**Supplementary Appendix 1: Model Specifications and Details**

Agent-based models (ABMs) are a class of computational models that simulate interactions of individual autonomous agents from which the system as a whole is evaluated. In most epidemiologic applications of ABMs, agents represent persons who are characterized by biological functions, positioning among other agents, exposures and disease states. Then, the model outputs are used to evaluate population-level outcomes. TITAN (1) was adapted to simulate an idealized two-stage randomized trial evaluating effect modification by component-level factors of the efficacy of randomized pre-exposure prophylaxis (PrEP) assignment among components of agents in a sexual network of men who have sex with men (MSM) in the city of Atlanta, Georgia (2, 3). The model described in this supplement was adapted from an existing ABM evaluating the impact of PrEP use on HIV incidence among MSM in Atlanta, while considering racial disparities (4-6).

**Supplementary Appendix 2: Parameter Data Sources and Agent Population**

To determine parameters for the model, empirical estimates specific to the population of MSM in Atlanta from the primary literature were used, if available. If those were not available, empirical estimates from Atlanta, or the closest match for geographic area, were employed. The population size was determined from published estimates for the geographic regions that contain Atlanta (4, 5) and published literature, as well as surveys, were used to inform the parameter estimates (references as specified in the Supplementary Table S1) (7). The estimated number of African American (AA) and non-Hispanic White (W) adult men (ages 18 to 74 years) in Atlanta was obtained from the American Community Survey (2012-2016, 5-year estimates) (8, 9). These proportions were then used with the estimated population of adult men ($n$= 154,832) to estimate the number of MSM. Based on the average of population estimates, the estimated population size was 11,245 MSM (7). The distribution of age was based upon estimates from the American Community Survey (2016 cycle, 5-year estimates) (8, 9). In 2016, the median ages in each of these populations were 32.3 years and 35.4 years, respectively (10, 11). The distribution of race and ethnicity was based on the distribution from the American Community Survey (2016 cycle, 5-year estimates) (8, 9). In 2016, there were an estimated 79,314 AA adult males (51.2%) and 75,518 W adult males (48.8%) (8, 9). Agents representing adult males of other racial/ethnic backgrounds were not considered, as these individuals represented a smaller proportion of adult males in this setting.

Given the two years of follow-up and the age distribution of the population, we assumed that there was no mortality and complete retention during follow-up. These assumptions were necessary because the existing methods for evaluating spillover do not allow for time-varying components (or spillover sets) in the sexual network. Several key parameters were stratified by race, including HIV prevalence, and engagement with the HIV treatment cascade. Prevalence of any drug use was also stratified by race (30% among African Americans, 49% among whites) (12-14). In addition, pre-enrollment HIV prevalence was also stratified by race (43% among African Americans, 13% among white) (15). Proportion with an HIV diagnosis, on ART and viral suppression among those on ART was also stratified by race (Supplementary Table S1).

At model initialization, agents were assigned an HIV status. HIV-infected agents were assigned the following: awareness of their status, utilization of antiretroviral treatment (ART), and achievement of viral suppression. Only HIV-negative agents were eligible to be randomized to PrEP in the simulated trial. We scaled the population size for this study to 17,440 agents to be higher than the estimated population size so that there would be more partnered agents (rather than just singletons) to better assess the spillover effects in the simulated trial.

An agent’s race and drug use status affected their sexual partnering preferences. Any drug use was defined as self-report of use of cannabis, cocaine, amphetamines, methamphetamines, inhalant nitrites, heroin/opioids, or benzodiazepines in the past 12 months (14). The empirical values for these race-specific parameters were derived from a prospective cohort designed to examine the individual, dyadic, and community-level factors that may contribute to the disparities in HIV prevalence and incidence between African American men who have sex with men (AAMSM) and non-Hispanic White men who have sex with men (WMSM) in Atlanta, Georgia (12). In this study, participants provided information regarding their five most recent sexual partners. AAMSM reported race concordance with their partners in 80.5% of dyads, whereas WMSM reported race concordance with their partners in 73.7% of dyads, which we used in our model parameters (12). For drug use, we assumed that 20% of substance using agents partnered only with other substance using agents.

All agents were assigned a sexual position preference (exclusively insertive, exclusively receptive, and versatile) at model initialization. The model included an absolute prohibition, such that two exclusively insertive agents could not pair with one another nor could two exclusively receptive agents. These parameters are derived from reports of role in last sex for up to 5 recent sexual partners as ascertained from the Involve[MEN]t cohort (15).

**Supplementary Appendix 3: Network Structure and Sexual Partnership Formation**

At model initialization for each model run, agents were assigned a target number of sexual partners (16-18). The total number of sexual partners per year was assumed to follow a negative binomial distribution with mean = 5 for African Americans and mean = 7 for whites (12). A target partner number was determined based on the total number of target partners and average partnership duration. The negative binomial is an appropriate distribution due overdispersion of the distribution of number of sexual partnerships in this population. The number of sexual acts per month within a partnership was assumed to follow a Poisson distribution, and each agent was assigned a total number of sexual acts per interval, based on a distribution of monthly number of sexual acts (15). For each agent, a pool of potential partners was created from all other agents seeking partners, and subsequently narrowed by sexual position and the agent’s assorting probabilities. The agent then selects partners from this generated pool to achieve its target number of partners.

Partnering occurred at model initiation, prior to trial enrollment, and the sociometric network remained static at all subsequent timepoints in the simulated trial. These partnerships at enrollment were used to determine network components with at least three agents. A network component is a subset of the agents of a network that are all connected through at least one partnership and not connected to any other agents in the network. Once formed, relationships were *not* dissolved and new relationships were *not* formed, but rather sexual networks were to be static, as ascertained prior to enrollment in the simulated trial (19).

**Supplementary Appendix 4: HIV Transmission**

Any two agents sharing a partnership (or edge) in the network could engage in anal intercourse. Sex acts using condoms were considered non-transmitting acts and therefore had no probability of HIV transmission. For each sexual act, condom use was a function of the number of previous contacts with that partner, where the likelihood of condom use decreases with more contact between two agents (20){Rosenberger, 2012 #3}. The base probabilities of condom use were informed by a cross-sectional assessment of condom use experiences as ascertained in the Involve[MEN]t cohort (21). For partnerships with condomless sex, there was a non-zero per-act probabilities of HIV transmission, which depended on position preference, PrEP (for HIV-negative agents), and adherence to PrEP at model initiation (for HIV-negative agents randomized to PrEP) (22). In addition, HIV-positive agents’ knowledge of their HIV status, ART utilization, and adherence to ART at model initiation also modified the per-act probability of HIV transmission (23). Given the low prevalence of injection risk behavior in this population, needle-sharing acts of injection drug use were not modeled.

**Supplementary Appendix 5: HIV Testing, Treatment and Disease Progression**

At each monthly interval, agents had a certain number of sex acts with their partners identified at model initiation. In the absence of any HIV intervention (PrEP or ART for treatment as prevention), any condomless sex act in a serodiscordant partnership had a non-zero probability of HIV transmission (per-act probability for condomless receptive anal intercourse was 1.38% and condomless insertive was 0.11%) (22).

Based on (23), we assumed that 65.5% of African American MSM and 81.8% of White MSM with HIV were diagnosed at model initialization. Agents aware of their infection status were assumed to have a lower transmission risk to their HIV-uninfected partners through the use of a scaling factor informed by research suggesting that newly-diagnosed MSM may decrease their numbers of sexual partners or avoid condomless anal intercourse with HIV-uninfected partners for a period of time following diagnosis (24). Probabilities of condom non-use within serodiscordant partnerships for which the HIV-infected agents were aware of their infection status were scaled by a factor of 0.5 (i.e., condom non-use was 50% less likely in these relationships).

Based on the Involve[MEN]t cohort, we assumed that 89.2% of AAMSM and 94.8% of all WMSM were ever tested for HIV infection. In subsequent time steps, 66.3% of AAMSM and 73.6% of WMSM were tested each calendar year. AAMSM obtained testing randomly with a monthly probability of 0.055 and WMSM obtained testing randomly with a monthly probability of 0.061. HIV-infected agents unaware of their infection tested at the same probability at HIV-uninfected agents. No restriction on re-testing within a year was imposed (i.e., an agent may test multiple times per year).

Once diagnosed, HIV-infected agents may initiate ART and become virally suppressed. Based on (23, 25), we assumed that 65.8% of AAMSM and 59.7% of WMSM who were diagnosed then initiated ART (representing 43.1% of all African American agents with HIV infection and 48.8% of all White agents with HIV infection). At model initialization, the proportion of diagnosed HIV-infected agents achieving optimal adherence (i.e., taking 90% or more of all doses) to ART were informed by the HIV care continuum surveillance activities. We assumed that 81.7% of white agents were fully adherent to ART (43.2% of all white agents with HIV infection), and 88.5% of African American agents were fully adherent to ART (35.2% of all African American agents with HIV infection) (23). We assumed adherence to ART was constant once agents initiate therapy. For viral suppression, we assumed that agents who have optimal adherence to ART will have viral load < 200 copies/mL.

The probability of HIV transmission varied depending upon the following factors: the HIV-negative agent’s PrEP use and adherence (assigned at the start of the simulated trial); if the HIV-infected agent knew their status; and the HIV-positive agent’s use of ART and subsequently, if they achieved viral suppression. For example, there was a 96% reduction in the risk of HIV acquisition for agents fully adherent to PrEP and a 76% reduction for those with less than full adherence (26). Once the per-act probability of transmission in a relationship was determined, the overall risk for the timestep was calculated and used to determine whether HIV transmission occurs in the partnership. Agents using HAART but not in viral suppression had 3% of the likelihood of HIV transmission. Agents that were virally suppressed had no ability to transmit; however, this model does not have risk reduction for diagnosed agents.

For progression to AIDS, a probability was assigned to all HIV-infected agents. Agents on ART were less likely to progress to AIDS than other HIV-infected agents. This was achieved through a scalar reduction in progression probability, with the reduction dependent on ART adherence. A base probability of progression of AIDS was assigned to all HIV-infected agents, where HIV-infected agents progressed to AIDS with a monthly probability of 0.51% (27-29). HIV-infected agent with optimal adherence to ART experienced AIDS progression with a monthly probability of 0.08%, which is a scalar reduction in this monthly probability of progression to AIDS by a factor of 6.375 (30-32). HIV-infected agents with suboptimal adherence to ART had identical probabilities of progression to AIDS as all other HIV-infected agents (regardless of their diagnosis status).

**Supplementary Appendix 6: Pre-Exposure Prophylaxis Use and Clinical Care**

In our simulated two-stage randomized trial, components within the sexual (sociometric) network were defined prior to enrollment in the trial among all MSM (ages 18 to 65 years); however, only components with at least one HIV-positive agent and three or more agents were eligible to become “intervention components,” and at the individual level, only HIV-negative agents with one or more partners were eligible to be enrolled in the trial and possibly randomized to PrEP. Depending on the allocation strategy assigned to the component, eligible agents were randomized to PrEP (or no PrEP). After enrollment in the trial, eligible agents who were randomized to the PrEP intervention received a 90-day supply, and then received a subsequent 90-day supply at each scheduled follow-up visit every 3 months. For the two-year duration of this simulated trial, all agents were retained in the study. After enrollment in the trial, agents were classified as adherent (defined as 4 or more doses per week) or suboptimal (defined as 2 to 4 doses per week). In the main model, 91.1% of WMSM and 56.8% of AAMSM were considered to be optimally adherent to PrEP at baseline (i.e., 4 or more doses per week) (33-35). Those with optimal adherence had a 96% reduction in the per-act probability of HIV acquisition, while those with partial adherence had a 76% reduction (26). This remained stable for an individual agent across all model runs.

**Supplementary Appendix 7: Model Calibration**

We used an iterative indirect approach following published guidelines (36). Outputs from the initial model setting were obtained and compared to surveillance data from the Georgia Department of Health (37). The model was then refined by adjusting parameters that had more known uncertainty in their values (e.g., monthly probability of HIV testing, frequency of engagement in condomless anal intercourse) with the goal of reducing the difference between the model output and available empirical data. We employed an iterative stepwise sweep of “calibration parameters” to align with observed empirical observations. Although this does not ensure the model is valid, this process assess if the model output aligns with empirical estimates (38). The main calibration target was annual HIV incidence by race based on a prospective cohort study of Black and White MSM in Atlanta (6.5 and 1.7 per 100 person-years, respectively (12). The model was able to reproduce these values as 6.37 (95% SI: 6.09, 6.58) and 1.61 (95% SI: 148, 172) per 100 person-years, respectively, for the scenario with no PrEP exposure. The calibration was accomplished by scaling parameters representing the number of sexual acts engaged per time step, race-based assortative missing in partnerships, and condom use. We then evaluated trajectories of diagnosed HIV infections, AIDS prevalence, and HAART prevalence among those diagnosed, and adjusted scalars on the monthly probabilities of testing for HIV infection, progression to AIDS, and ART enrollment/discontinuation to ensure these values remained stable about the parameterized initial condition at the start of the model run. Complete details and results of the calibration procedure for this model have been previously published (4, 5).

**Supplementary Appendix 8: Technical Details**

Python software, version 2.7.12 (39), along with the NumPy (40) and NetworkX (41) packages, was used for coding, testing, and performing sensitivity analyses of this model. The analysis of model output for this paper was generated using SAS software (version 9.4). Copyright © 2021 SAS Institute Inc. SAS and all other SAS Institute Inc. product or service names are registered trademarks or trademarks of SAS Institute Inc., Cary, NC, USA. R software, version 3.5.1 (R Foundation for Statistical Computing, Vienna, Austria) (42), along with ggplot2 (43), was used to produce figures. At each monthly time interval, variable information on each agent and their component members were recorded, including HIV status, awareness of status, utilization of ART and viral suppression (if HIV positive), and number of sexual acts. Agents were assigned characteristics at model initiation and components were determined prior to enrollment in the simulated trial based on sexual partnerships formed at model initialization.

The simulations were run on Oscar, Brown University’s supercomputing cluster. Oscar operates on the CentOS 6.7 Linux operating system and utilizes the SLURM workload manager. The simulations were processed using 2.53 GHz Intel Xeon E5540 processors operating with 8 cores at 14.84 teraflops and 12GB of DDR3 memory. The model was run for 24 monthly time steps (2 years), included 17,440 agents total per model run, and averages over a total of 1,000 Monte Carlo runs per scenario, each with a simulated population based on the model parameters described above.

**Supplementary Appendix 9: Sensitivity Analyses for Intervention Coverage and Estimators**

For the sensitivity analysis, we simulated a trial with 30% coverage of PrEP in the intervention components, as well as the 70% coverage scenario. The cumulative incidence of HIV over 24 months in the simulated trial and stratified by each effect modifier are displayed by intervention and control components in Supplementary Table S2. We estimated the spillover effects of PrEP on HIV incidence by each effect modifier using individual- and component-weighted estimators (both stabilized and unstabilized) for the risk difference and risk ratio (Supplementary Tables S3 to S6). In general, the estimated spillover effects were larger in magnitude with 70% intervention coverage, as compared to 30% coverage. The results for the stabilized estimators were comparable to the unstabilized; however, there were some slight differences between the individual-weighted and component-weighted estimators, which is expected as the sizes of the components varied in the simulated trials. Interestingly, the trends observed in the main analysis (e.g., larger in magnitude estimated spillover effects among components with a lower prevalence of drug use) remained in this sensitivity analysis.

**Supplementary Appendix 10: One-Way Sensitivity Analyses**

We performed one-way sensitivity analyses to assess the impact of our model parameterization on model results for HIV cumulative incidence. We focused our reporting on two-stage randomized trials with 70% coverage allocation strategies in the intervention components because this was our primary model scenario. We used the component-weighted stabilized estimators in this sensitivity analysis. This sensitivity analysis evaluated the model results while varying assumptions about adherence to PrEP and discontinuation of PrEP. In the main model, 91.1% adherence for WMSM and 56.8% for AAMSM were considered to be optimally adherent to PrEP at baseline (i.e., 4 or more doses per week). In the sensitivity analysis, we considered 80% PrEP adherence for WMSM and 50% PrEP adherence for AAMSM. In the main model, no agents discontinued PrEP during follow-up, and in the sensitivity analysis, 10% of agents discontinued PrEP in each monthly interval (44).

In Supplementary Tables S7 to S10, we display the HIV prevalence and HIV cumulative incidence at the end of two years of follow-up after randomization based on a simulated trial with 70% coverage with 80% PrEP adherence for WMSM and 50% PrEP adherence for AAMSM. In most cases, the sensitivity analysis resulted in slightly more agents on PrEP with incident HIV, while also modest increases in HIV incidence among agents in intervention components but not on PrEP. In Supplementary Tables S11 to S14, we display the HIV prevalence and HIV cumulative incidence at the end of two years of follow-up after randomization based on a simulated trial with 70% coverage with 10% discontinuation of PrEP. In most cases, the sensitivity analysis resulted in substantially more agents on PrEP with incident HIV, while also small or no changes HIV incidence among agents in intervention components but not on PrEP.

Supplementary Table S15 to S18 display the estimated spillover risk differences (RD) and risk ratios (RR) for HIV cumulative incidence in a two-stage randomized trial with 70% coverage. In the main analysis, 91.1% adherence for WMSM and 56.8% for AAMSM were considered to be optimally adherent to PrEP at baseline (i.e., 4 or more doses per week). In the sensitivity analysis, we considered 80% PrEP adherence for WMSM and 50% PrEP adherence for AAMSM. Interestingly, the estimated spillover effects were fairly robust to this one-way sensitivity analysis for PrEP adherence and the results were comparable to the main analysis. Supplementary Figure S1 and Supplementary Figure S2 display the estimated spillover effects when PrEP adherence was lower in a simulated trial with 70% PrEP coverage on the difference and ratio, respectively.

Supplementary Table S15 to S18 display the estimated spillover risk differences (RD) and risk ratios (RR) for HIV cumulative incidence in a two-stage randomized trial with 70% coverage. In the main analysis, we assumed that no agents discontinued PrEP. In the sensitivity analysis, we considered a scenario where 10% of agents discontinued PrEP. Interestingly and despite increases in HIV incidence among those on PrEP, the estimated spillover effects were fairly robust to this one-way sensitivity analysis for PrEP discontinuation and the results were comparable to the main analysis. Supplementary Figure S3 and Supplementary Figure S4 display the estimated spillover effects when PrEP discontinuation was higher (10%) in a simulated trial with 70% PrEP coverage on the difference and ratio, respectively.

**Supplementary Appendix 11: Causal Inference Methods for Evaluation of Effect Modification in the Presence of Spillover in an ABM**

In each simulation, we employed two-stage randomization, which is a randomized trial design that allows for quantifying intervention effects in the presence of spillover (i.e., interference or dissemination). Spillover is when one agent’s intervention assignment affects another agent’s outcome. In a two-stage randomized design, components are first randomized to the intervention (e.g., 70% coverage of PrEP) or control (e.g., 0% coverage of PrEP), then according to the component-level allocation strategy, agents are randomly assigned to the intervention. In each run of the model, the component sizes vary, so we extended estimators from Basse and Feller (2018) to consider effect modification by component characteristics (45, 46). We assumed partial and stratified interference. That is, we assume that the intervention assignment influences others in the same component only; however, this influence does not extend beyond the component (47). We also assume *stratified interference* where an individual’s potential outcome is dependent only on their own intervention assignment and the proportion of agents exposed in the component (48). We also make the usual assumptions required for causal inference (exchangeability, consistency, and positivity) (49-52). We assume a Bernoulli allocation strategy for intervention assignment within each intervention component (48).

There are $I$ components total and each of the component has $n_{i}$ individuals for $j=1,2,...,n_{i}$ and $\sum_{i=1}^{I} n_{i}=N$. Let $Y_{ij}, A_{ij}$ represent an observed outcome and treatment assignment status of $j^{th}$ agent in component $i$. Let $C_{i}$be an indicator for the treatment assignment at the component level. Let $M_{i}$ be a component-level variable measured at baseline (e.g., covariates aggregated to the component level or network characteristic of the component). We estimated the spillover effect within levels of the following component-level effect modifiers aggregated to the component level: HIV prevalence and drug use prevalence; and network characteristics: average density and average bridging potential (53-55). For now, we consider only categorical versions of the effect modifier $M_{i}$. We consider the potential outcome for agent $j$ in component $i$ as $Y_{ij}(\boldsymbol{A}_{i})=Y_{ij}(C_{i}=c, A_{ij}=a)$. Because we have a control group with no agents assigned to PrEP, there are three possible combinations of the intervention resulting in three potential outcomes: $Y_{ij}(1,1), Y_{ij}(1,0), Y_{ij}(0,0)$. The observed outcome is a function of the intervention assignment and potential outcomes; that is, $Y_{ij}^{obs}=C_{i}A_{ij}Y_{ij}\left( 1,1 \right)+C_{i}\left( 1-A_{ij} \right)Y_{ij}\left( 1,0 \right)+(1-C_{i})Y_{ij}(0,0).$Let $T_{ca}=\{(i,j): C_{i}=c and A_{ij}=a\}$to denote the set of components and agents who are assigned to $C_{i}=c$ and $A_{ij}=a$.

In the setting with varying component sizes, there are two types of estimands: component-weighted estimands that assign equal weight to components, regardless of the number of individuals in each component; and individual-weighted estimands that assign equal weight to individuals, regardless of the distribution across components (45). The spillover (i.e., disseminated or indirect) effect is $IE=\sum_{i=1}^{I} {w^{*}}_{i}\sum_{j=1}^{n_{i}} [Y_{ij}\left( 1,0 \right)-Y_{ij}\left( 0,0 \right)]$, where ${w^{*}}_{i}=\frac{1}{In_{i}}$ corresponds to component-weighted estimands and ${w^{*}}_{i}=\frac{1}{N}$corresponds to individual-weighted estimands with $N=\sum_{i} n_{i}$.

To estimate the spillover effect, we employ the two-stage inverse probability weights ${w_{i}}^{(00)}$, ${w_{i}}^{(10)}$, ${w_{i}}^{(11)}$ as ${w_{i}}^{(11)}=\frac{1}{Pr(C_{i}=1)}\frac{1}{Pr({A_{ij}=1|C}_{i}=1)}$, ${w_{i}}^{(10)}=\frac{1}{Pr(C_{i}=1)}\frac{1}{Pr({A_{ij}=0|C}_{i}=1)}$, and ${w_{i}}^{(0)}=\frac{1}{Pr(C_{i}=0)}$. Also define ${w_{i}}^{(c)}=\frac{1}{Pr(C_{i}=c )}$. The weighted spillover effect estimator is:

$\hat{IE}=\sum_{(i,j)\in T_{10}} {{w_{i}^{*}w}_{i}}^{(10)}{Y^{obs}}_{ij}(1,0)-\sum_{(i,j)\in T_{00}} {{w_{i}^{*}w}_{i}}^{\left( 00 \right)}{Y^{obs}}_{ij}(0,0)$.

These are unbiased estimators in a two-stage randomized design (45).

We define the effect measure modification parameters as follows, modestly extending results in (32). We define these on the difference scale below, but will also consider both the relative and absolute scales when analyzing the model results. Let the indicator function $I\left( M_{i}=m \right)=1$ if $M_{i}=1$ and $I\left( M_{i}=m \right)=0$, otherwise. Let $I_{m}$denote the number of components with $M_{i}=m$ and $n_{m}^{+}=\sum_{i} n_{i}I(M_{i}=m)$ denote the number of individuals in stratum $M_{i}=m$. Among components with $M_{i}=m$, the spillover (i.e., indirect) effect is

$IE_{m}=\sum_{i=1}^{I} {{I\left( M_{i}=m \right)w}^{*}}_{mi}\sum_{j=1}^{n_{i}} [Y_{ij}\left( 1,0 \right)-Y_{ij}\left( 0,0 \right)]$,

where ${w^{*}}_{mi}=\frac{1}{I_{m}n_{i}}$ corresponds to component-weighted estimands and ${w^{*}}_{mi}=\frac{1}{n_{m}^{+}}$corresponds to individual-weighted estimands.

To quantify the spillover effect modified by $M$, we proposed the following modifications to the two-stage inverse probability weights: ${w_{i}}^{(m11)}=\frac{1}{Pr(C_{i}=1|M_{i}=m)}\frac{1}{Pr({A_{ij}=1|C}_{i}=1,M_{i}=m)}$, ${w_{i}}^{(m10)}=\frac{1}{Pr(C_{i}=1{|M}_{i}=m)}\frac{1}{Pr({A_{ij}=0|C}_{i}=1,M_{i}=m)}$, and ${w_{i}}^{(m00)}=\frac{1}{Pr(C_{i}=0|M_{i}=m)}$. Also define ${w_{i}}^{(mc)}=\frac{1}{Pr(C_{i}=c |M_{i}=m)}$. We also revised the individual and component level weights: ${w^{*}}_{mi}=\frac{1}{I_{m}n_{i}}$ corresponds to component-weighted estimands with and ${w^{*}}_{mi}=\frac{1}{n_{m}^{+}}$corresponds to individual-weighted estimands. Within each level of $M,$ the weighted spillover effect estimator is

$$\hat{IE}_{m}=\sum_{(i,j)\in T_{10}} {{{{I(M}_{i}=m)w}_{mi}^{*}w}_{i}}^{(m10)}{Y^{obs}}_{ij}(1,0)-\sum_{\left( i,j \right)\in T_{00}} {{{{I(M}_{i}=m)w}_{mi}^{*}w}_{i}}^{\left( m00 \right)}{Y^{obs}}_{ij}\left( 0,0 \right).$$

This estimator is unbiased in a two-stage randomized design with a single allocation strategy and a control group (no agents were assigned to PrEP) (45, 56). The estimators of the risk ratio of the spillover effect is defined analogously and can be estimated similarly to the risk difference.

We also consider stabilized versions of the weights and proposed Hajek-type estimators, which may provide more efficient estimators in a simulation setting and are a modest extension of existing estimators. We extended an expression for the spillover effect estimator and follow the proposed stabilized estimators described in (45). Let $\theta_{i}^{(10)}=\frac{1}{Pr\left( C_{i}=1 \right)Pr(A_{ij}=0)}$ and $\theta_{i}^{(00)}=\frac{1}{Pr(C_{i}=0)}$, then define:

$$\theta_{+}^{(10)}= \sum_{i} I\left( C_{i}=1 \right)\sum_{j} I\left( A_{ij}=0 \right)\theta_{i}^{(10)}$$

$$\theta_{+}^{(00)}=\sum_{i} I\left( C_{i}=0 \right)\sum_{j} I(A_{ij}=0)\theta_{i}^{(00)}$$

Let $I$ be the total number of components, $I_{1}$ be the number of intervention components, and $I_{0}$ be the number of control components. Let $n^{+}=\sum_{i} n_{i}$, $n_{00}^{+}= \sum_{i} I\left( C_{i}=0 \right)\sum_{j} I(A_{ij}=0)$ and $n_{10}^{+}= \sum_{i} I\left( C_{i}=1 \right)\sum_{j} I(A_{ij}=0)$. Then, $\theta_{i}^{(00)}=\frac{I}{I_{0}}$ and so $\theta_{+}^{(00)}=\frac{I}{I_{0}}n_{00}^{+}$. Also, $\theta_{i}^{(10)}=\frac{I}{I_{1}}\frac{1}{Pr(A_{ij}=0)}$ and so $\theta_{+}^{(10)}=n_{10}^{+}\frac{I}{I_{1}}\frac{1}{Pr(A_{ij}=0)}$. Then, with a control group with no agents randomized to the intervention,

$$E\left( \theta_{+}^{\left( 00 \right)} \right)=\frac{I}{I_{0}}\frac{I_{0}}{I}\sum_{i} n_{i}=n^{+}$$

$$E\left( \theta_{+}^{\left( 10 \right)} \right)=\frac{I}{I_{1}}\frac{I_{1}}{I}\sum_{i} \sum_{j} \frac{\Pr\left( A_{ij}=0 \right)}{Pr(A_{ij}=0)}=\sum_{i} n_{i}=n^{+}$$

Their proposed spillover effect estimator with stabilized weights is (33):

$$\hat{IE}=\frac{E\left( \theta_{+}^{\left( 10 \right)} \right)}{\theta_{+}^{\left( 10 \right)}}\sum_{i} \sum_{j} I\left( C_{i}=1 \right)I\left( A_{ij}=0 \right)\theta_{i}^{\left( 10 \right)}w_{i}^{*}Y_{ij}\left( 1,0 \right)- \frac{E\left( \theta_{+}^{\left( 00 \right)} \right)}{\theta_{+}^{\left( 00 \right)}}\sum_{i} \sum_{j} I\left( C_{i}=0 \right)\theta_{i}^{\left( 00 \right)}w_{i}^{*}Y_{ij}\left( 0,0 \right)$$

$=\frac{n^{+}}{n_{10}^{+}\frac{I}{I_{1}}\frac{1}{Pr(A_{ij}=0)}}\sum_{i} \sum_{j} I\left( C_{i}=1 \right)I\left( A_{ij}=0 \right)\theta_{i}^{\left( 10 \right)}w_{i}^{*}Y_{ij}\left( 1,0 \right)- \frac{n^{+}}{n_{00}^{+}\frac{I}{I_{0}}}\sum_{i} \sum_{j} I\left( C_{i}=0 \right)\theta_{i}^{\left( 00 \right)}w_{i}^{*}Y_{ij}\left( 0,0 \right)$.

When considering effect modification, we can estimate effects separately among the stratum determined by levels of $M$. Let $I_{m}$ be the total number of components with $M_{i}=m$, $I_{1m}$ be the number of intervention components with $M_{i}=m$, and $I_{0m}$ be the number of control components with $M_{i}=m$. Recall $n_{m}^{+}=\sum_{i} n_{i}I(M_{i}=m)$, and let $n_{m00}^{+}= \sum_{i} I(M_{i}=m)I\left( C_{i}=0 \right)\sum_{j} I(A_{ij}=0)$ and $n_{m10}^{+}= \sum_{i} I(M_{i}=m)I\left( C_{i}=1 \right)\sum_{j} I(A_{ij}=0)$. Then, $\theta_{mi}^{(m00)}=\frac{I_{m}}{I_{0m}}$ and so $\theta_{m+}^{(m00)}=\frac{I_{m}}{I_{0m}}n_{m00}^{+}$. Also, $\theta_{mi}^{(m10)}=\frac{I_{m}}{I_{1m}}\frac{1}{Pr(A_{ij}=0)}$ and so $\theta_{m+}^{(m10)}=n_{m10}^{+}\frac{I_{m}}{I_{1m}}\frac{1}{Pr(A_{ij}=0)}$. Then, the stabilized estimator of the spillover effect among components with $M_{i}=m$is:

$\hat{IE}_{m}=\frac{n_{m}^{+}}{n_{m10}^{+}\frac{I_{m}}{I_{1m}}\frac{1}{Pr(A_{ij}=0)}}\sum_{i} \sum_{j} I\left( M_{i}=m \right)I\left( C_{i}=1 \right)I\left( A_{ij}=0 \right)\theta_{i}^{\left( m10 \right)}w_{mi}^{*}Y_{ij}\left( 1,0 \right)- \frac{\sum_{i} \sum_{j} Pr(A_{ij}=0)}{n_{m00}^{+}\frac{I_{m}}{I_{0m}}}\sum_{i} \sum_{j} I\left( M_{i}=m \right)I\left( C_{i}=0 \right)\theta_{i}^{\left( m00 \right)}w_{mi}^{*}Y_{ij}\left( 0,0 \right)$.

**Supplementary Table S1.** Summary of key model parameters in an agent-based model simulating a two-stage randomized trial overall and in a population of AAMSM and WMSM

| **Domain** | **Overall** | **AAMSM** | **WMSM** | **Source** |
| --- | --- | --- | --- | --- |
| **Demographic Characteristics** |  |  |  |  |
| Population size (*n*) | 17,440 | 6,784 | 10,656 | (57) |
| Age distribution (%) |  |  |  | (57) |
| 18 to 24 years |  | 18.0 | 15.1 |  |
| 25 to 34 years |  | 22.6 | 28.2 |  |
| 35 to 44 years |  | 18.4 | 21.0 |  |
| 45 to 54 years |  | 16.9 | 17.3 |  |
| 55 to 64 years |  | 14.3 | 11.6 |  |
| 65 years |  | 9.9 | 6.7 |  |
| Drug use |  | 30.0 | 48.5 | (12, 13) |
| **Sexual Behaviors** |  |  |  |  |
| Number of sex partners per year (%) |  |  |  | (16) |
| Median (Interquartile Range) |  | 5 (3-9) | 7 (4-12) |  |
| Probability of condom use |  |  |  |  |
| 0 prior encounters with partner |  | 0.688 | 0.528 | (20) |
| 1 prior encounter with partner |  | 0.629 | 0.483 |  |
| 2 to 9 prior encounters with partner |  | 0.578 | 0.444 |  |
| ≥10 prior encounters with partner |  | 0.198 | 0.152 |  |
| Probability of HIV transmission (per act) |  |  |  |  |
| Condomless insertive anal intercourse | 0.0011 |  |  | (22) |
| Condomless receptive anal intercourse | 0.0138 |  |  | (22) |
| Sexual Role |  |  |  |  |
| Insertive only (%) |  | 24.2 | 22.8 | (15) |
| Receptive only (%) |  | 32.1 | 22.8 | (15) |
| Versatile (%) |  | 43.7 | 54.4 | (15) |
| **Pre-Exposure Prophylaxis Use** |  |  |  |  |
| Retention in clinical care, 6 months (%) | 100.0 |  |  | (33) |
| Optimal adherence (≥4 pills per week) (%) |  | 91.1 | 56.8 | (33) |
| Reduction in risk of HIV acquisition (%) |  |  |  | (26) |
| Optimal adherence | 96.0 |  |  |  |
| Suboptimal adherence | 76.0 |  |  |  |
| **HIV and Cascade of Care** |  |  |  |  |
| HIV prevalence (% of all MSM) |  | 43.4 | 13.2 | (23) |
| HIV diagnosed (% of HIV+) |  | 65.5 | 81.8 | (23) |
| ART utilization (% of HIV+) |  | 43.1 | 48.8 | (23) |
| Virologically suppressed (% of HIV+) |  | 35.2 | 43.2 | (23) |
| **Assortative Mixing** |  |  |  |  |
| WMSM with WMSM | 73.7 |  |  | (12) |
| AAMSM with AAMSM | 80.5 |  |  | (12) |
| Substance using agents with  Substance using agents | 20.0 |  |  |  |

Abbreviation: men who have sex with men (MSM); African American MSM (AAMSM); White MSM (WMSM).

**Supplementary Table S2.** Cumulative incidence of HIV over two years of follow-up after two-stage randomization stratified by four modifiers among agents within PrEP intervention (30% coverage) and control components in an agent-based model representing among men who have sex with men Atlanta, Georgia, 2015-2017 (n = 3,947)

|  | Intervention Components | | | | | | Control Components | | |
| --- | --- | --- | --- | --- | --- | --- | --- | --- | --- |
|  | Agents on PrEP | | | Agents Not on PrEP | | | Agents Not on PrEP | | |
| Effect Modifiers | Total Agents | HIV+ | Cumulative Incidence | Total Agents | HIV+ | Cumulative Incidence | Total Agents | HIV+ | Cumulative Incidence |
| Overall | 386.3 | 4.7 | 0.01 | 900.5 | 84.5 | 0.09 | 1287.4 | 124.1 | 0.10 |
| Drug use |  |  |  |  |  |  |  |  |  |
| Among $M = 0$ | 208.9 | 2.9 | 0.01 | 487.1 | 50.4 | 0.10 | 695.3 | 73.8 | 0.11 |
| Among $M = 1$ | 177.4 | 1.8 | 0.01 | 413.4 | 34.1 | 0.08 | 592.1 | 50.4 | 0.09 |
| HIV Prevalence |  |  |  |  |  |  |  |  |  |
| Among $M = 0$ | 297.9 | 2.8 | 0.01 | 694.2 | 52.3 | 0.08 | 992.7 | 77.2 | 0.08 |
| Among $M = 1$ | 88.5 | 2.0 | 0.02 | 206.3 | 32.2 | 0.16 | 294.8 | 46.9 | 0.16 |
| Bridging Potential |  |  |  |  |  |  |  |  |  |
| Among $M = 0$ | 149.9 | 2.1 | 0.01 | 349.0 | 39.1 | 0.11 | 497.4 | 56.8 | 0.11 |
| Among $M = 1$ | 236.4 | 2.6 | 0.01 | 551.5 | 45.4 | 0.08 | 790.0 | 67.3 | 0.09 |
| Density |  |  |  |  |  |  |  |  |  |
| Among $M = 0$ | 305.0 | 3.5 | 0.01 | 712.5 | 61.9 | 0.09 | 1018.5 | 91.5 | 0.09 |
| Among $M = 1$ | 81.3 | 1.2 | 0.02 | 188.0 | 22.6 | 0.12 | 268.9 | 32.6 | 0.12 |

**Supplementary Table S3.** Spillover effects of PrEP on cumulative incidence of HIV over two years of follow-up stratified by modifier *drug use* (M=0 vs. M=1) after two-stage randomization among agents within PrEP intervention (30% coverage) and control components in an agent-based model representing men who have sex with men, Atlanta, Georgia, 2015-2017 (n = 3,947)^1^

|  | **Unstabilized** | | **Stabilized** | |
| --- | --- | --- | --- | --- |
| **Component Weighted** | RD (95% SI) | RR (95% SI) | RD (95% SI) | RR (95% SI) |
| M=0 |  |  |  |  |
| 30% Coverage | -0.03 (-0.09, 0.04) | 0.86 (0.52, 1.3) | -0.03 (-0.09, 0.04) | 0.86 (0.52, 1.3) |
| 70% Coverage | -0.09 (-0.14, -0.03) | 0.47 (0.24, 0.77) | -0.08 (-0.14, -0.03) | 0.47 (0.24, 0.78) |
| M=1 |  |  |  |  |
| 30% Coverage | -0.02 (-0.10, 0.06) | 0.90 (0.43, 1.6) | -0.02 (-0.10, 0.06) | 0.90 (0.43, 1.6) |
| 70% Coverage | -0.07 (-0.14, 0.00) | 0.53 (0.20, 1.0) | -0.07 (-0.14, 0.00) | 0.53 (0.20, 1.0) |
| **Individual Weighted** | RD (95% SI) | RR (95% SI) | RD (95% SI) | RR (95% SI) |
| M=0 |  |  |  |  |
| 30% Coverage | -0.02 (-0.07, 0.03) | 0.87 (0.54, 1.3) | -0.02 (-0.07, 0.03) | 0.87 (0.54, 1.3) |
| 70% Coverage | -0.07 (-0.11, -0.02) | 0.50 (0.27, 0.78) | -0.06 (-0.11, -0.02) | 0.50 (0.27, 0.78) |
| M=1 |  |  |  |  |
| 30% Coverage | -0.01 (-0.07, 0.04) | 0.91 (0.49, 1.5) | -0.01 (-0.07, 0.04) | 0.91 (0.49, 1.5) |
| 70% Coverage | -0.05 (-0.09, 0.00) | 0.57 (0.24, 1.0) | -0.05 (-0.09, 0.00) | 0.57 (0.25, 1.0) |

**Supplementary Table S4.** Spillover Effects of PrEP on cumulative incidence of HIV over two years of follow-up stratified by modifier *HIV prevalence* (M=0 vs. M=1) after two-stage randomization among agents within PrEP intervention (30% coverage) and control components with 95% simulation intervals (SI) in an agent-based model representing men who have sex with men, Atlanta, Georgia, 2015-2017 (n = 3,947)^1^

|  | Unstabilized | | Stabilized | |
| --- | --- | --- | --- | --- |
| **Component Weighted** | RD (95% SI) | RR (95% SI) | RD (95% SI) | RR (95% SI) |
| M=0 |  |  |  |  |
| 30% Coverage | -0.01 (-0.07, 0.04) | 0.92 (0.54, 1.5) | -0.01 (-0.07, 0.04) | 0.92 (0.54, 1.5) |
| 70% Coverage | -0.05 (-0.10, -0.01) | 0.56 (0.26, 0.91) | -0.05 (-0.10, -0.01) | 0.56 (0.26, 0.91) |
| M=1 |  |  |  |  |
| 30% Coverage | -0.05 (-0.16, 0.06) | 0.81 (0.43, 1.4) | -0.05 (-0.16, 0.06) | 0.81 (0.42, 1.4) |
| 70% Coverage | -0.13 (-0.22, -0.04) | 0.41 (0.18, 0.76) | -0.13 (-0.22, -0.04) | 0.41 (0.18, 0.76) |
| **Individual Weighted** | RD (95% SI) | RR (95% SI) | RD (95% SI) | RR (95% SI) |
| M=0 |  |  |  |  |
| 30% Coverage | -0.01 (-0.04, 0.03) | 0.93 (0.58, 1.4) | -0.01 (-0.04, 0.03) | 0.93 (0.59, 1.4) |
| 70% Coverage | -0.04 (-0.07, -0.01) | 0.60 (0.31, 0.94) | -0.04 (-0.07, -0.01) | 0.60 (0.31, 0.95) |
| M=1 |  |  |  |  |
| 30% Coverage | -0.03 (-0.12, 0.05) | 0.83 (0.47, 1.3) | -0.03 (-0.12, 0.05) | 0.84 (0.47, 1.3) |
| 70% Coverage | -0.10 (-0.17, -0.03) | 0.46 (0.21, 0.79) | -0.10 (-0.17, -0.03) | 0.46 (0.21, 0.80) |

**Supplementary Table S5.** Spillover effects of PrEP on cumulative incidence of HIV over two years of follow-up stratified by modifier *bridging potential* (M=0 vs. M=1) after two-stage randomization among agents within PrEP intervention (30% coverage) and control components with 95% simulation intervals (SI) in an agent-based model representing men who have sex with men, Atlanta, Georgia, 2015-2017 (n = 3,947)^1^

|  | Unstabilized | | Stabilized | |
| --- | --- | --- | --- | --- |
| **Component Weighted** | RD (95% SI) | RR (95% SI) | RD (95% SI) | RR (95% SI) |
| M=0 |  |  |  |  |
| 30% Coverage | -0.02 (-0.08, 0.03) | 0.84 (0.50, 1.3) | -0.02 (-0.08, 0.03) | 0.84 (0.50, 1.3) |
| 70% Coverage | -0.08 (-0.12, -0.03) | 0.44 (0.23, 0.72) | -0.08 (-0.12, -0.03) | 0.44 (0.23, 0.72) |
| M=1 |  |  |  |  |
| 30% Coverage | -0.01 (-0.07, 0.05) | 0.96 (0.53, 1.6) | -0.01 (-0.06, 0.05) | 0.96 (0.53, 1.6) |
| 70% Coverage | -0.04 (-0.09, 0.02) | 0.68 (0.31, 1.2) | -0.04 (-0.09, 0.02) | 0.68 (0.31, 1.2) |
| **Individual Weighted** | RD (95% SI) | RR (95% SI) | RD (95% SI) | RR (95% SI) |
| M=0 |  |  |  |  |
| 30% Coverage | -0.02 (-0.08, 0.03) | 0.85 (0.50, 1.3) | -0.02 (-0.08, 0.03) | 0.85 (0.50, 1.3) |
| 70% Coverage | -0.07 (-0.12, -0.03) | 0.44 (0.23, 0.71) | -0.07 (-0.12, -0.03) | 0.44 (0.23, 0.71) |
| M=1 |  |  |  |  |
| 30% Coverage | -0.01 (-0.05, 0.04) | 0.96 (0.56, 1.50) | -0.01 (0.05, 0.04) | 0.96 (0.56, 1.5) |
| 70% Coverage | -0.03 (-0.08, 0.02) | 0.70 (0.34, 1.21) | -0.03 (-0.07, 0.02) | 0.70 (0.34, 1.2) |

**Supplementary Table S6.** Spillover effects of PrEP on cumulative incidence of HIV over two years of follow-up stratified by modifier *Density* (M=0 vs. M=1) after two-stage randomization among agents within PrEP intervention (30% coverage) and control components with 95% simulation intervals (SI) in an agent-based model representing men who have sex with men, Atlanta, Georgia, 2015-2017 (n = 3,947)^1^

|  | Unstabilized | | Stabilized | |
| --- | --- | --- | --- | --- |
| **Component Weighted** | RD (95% SI) | RR (95% SI) | RD (95% SI) | RR (95% SI) |
| M=0 |  |  |  |  |
| 30% Coverage | -0.01 (-0.07, 0.04) | 0.91 (0.54, 1.4) | -0.01 (-0.07, 0.04) | 0.92 (0.54, 1.4) |
| 70% Coverage | -0.06 (-0.10, -0.01) | 0.58 (0.30, 0.94) | -0.06 (-0.10, -0.01) | 0.58 (0.30, 0.94) |
| M=1 |  |  |  |  |
| 30% Coverage | -0.03 (-0.09, 0.04) | 0.83 (0.45, 1.4) | -0.03 (-0.09, 0.04) | 0.84 (0.45, 1.4) |
| 70% Coverage | -0.08 (-0.14, -0.02) | 0.42 (0.17, 0.79) | -0.08 (-0.14, -0.02) | 0.42 (0.17, 0.79) |
| **Individual Weighted** | RD (95% SI) | RR (95% SI) | RD (95% SI) | RR (95% SI) |
| M=0 |  |  |  |  |
| 30% Coverage | -0.01 (-0.05, 0.03) | 0.92 (0.59, 1.3) | -0.01 (-0.05, 0.03) | 0.92 (0.59, 1.3) |
| 70% Coverage | -0.04 (-0.08, -0.01) | 0.61 (0.34, 0.93) | -0.04 (-0.08, -0.01) | 0.62 (0.34, 0.93) |
| M=1 |  |  |  |  |
| 30% Coverage | -0.03 (-0.09, 0.04) | 0.83 (0.45, 1.4) | -0.03 (-0.09, 0.04) | 0.84 (0.45, 1.4) |
| 70% Coverage | -0.08 (-0.14, -0.02) | 0.42 (0.17, 0.79) | -0.08 (-0.14, -0.02) | 0.42 (0.17, 0.79) |

**Supplementary Table S7.** Sensitivity analyses of PrEP adherence, HIV prevalence and incidence in a two-stage randomization with 70% coverage in the intervention group in an agent-based model representing among men who have sex with men Atlanta, Georgia, 2015-2017, stratified by the effect modifier $M$ (drug use).

| **Scenario** | **HIV Prevalence** | | | **Cumulative Incidence** | | |
| --- | --- | --- | --- | --- | --- | --- |
|  | **Intervention**  **Agents on PrEP** | **Intervention Agents not on PrEP** | **Control Agents** | **Intervention**  **Agents on PrEP** | **Intervention Agents not on PrEP** | **Control Agents** |
| **Among Components with *M* = 1** | | | | | | |
| Main | 4.0 | 291.8 | 327.4 | 4.0 | 14.3 | 51.0 |
| *PrEP Adherence* |  |  |  |  |  |  |
| 80% (White MSM)  50% (Black MSM) | 4.6 | 291.2 | 329.8 | 4.6 | 14.3 | 51.4 |
| **Among Components with *M* = 0** | | | | | | |
| Main | 6.4 | 421.1 | 471.6 | 6.4 | 21.1 | 74.2 |
| *PrEP Adherence* |  |  |  |  |  |  |
| 80% (White MSM)  50% (Black MSM) | 7.5 | 422.4 | 478.0 | 7.5 | 21.4 | 74.9 |

**Supplementary Table S8.** Sensitivity analyses of PrEP adherence, HIV prevalence and incidence in a two-stage randomization with 70% coverage in the intervention group in an agent-based model representing among men who have sex with men Atlanta, Georgia, 2015-2017, stratified by the effect modifier $M$ (HIV prevalence).

| **Scenario** | **HIV Prevalence** | | | **Cumulative Incidence** | | |
| --- | --- | --- | --- | --- | --- | --- |
|  | **Intervention**  **Agents on PrEP** | **Intervention Agents not on PrEP** | **Control Agents** | **Intervention**  **Agents on PrEP** | **Intervention Agents not on PrEP** | **Control Agents** |
| **Among Components with *M* = 1** | | | | | | |
| Main | 4.4 | 358.0 | 389.3 | 4.4 | 13.8 | 47.7 |
| *PrEP Adherence* |  |  |  |  |  |  |
| 80% (White MSM)  50% (Black MSM) | 5.1 | 358.4 | 395.0 | 5.1 | 14.2 | 48.5 |
| **Among Components with *M* = 0** | | | | | | |
| Main | 6.0 | 354.9 | 409.8 | 6.0 | 21.6 | 77.5 |
| *PrEP Adherence* |  |  |  |  |  |  |
| 80% (White MSM)  50% (Black MSM) | 7.1 | 355.2 | 412.8 | 7.1 | 21.6 | 77.9 |

**Supplementary Table S9.** Sensitivity analyses of PrEP adherence, HIV prevalence and incidence in a two-stage randomization with 70% coverage in the intervention group in an agent-based model representing among men who have sex with men Atlanta, Georgia, 2015-2017, stratified by the effect modifier $M$ (bridging potential).

| **Scenario** | **HIV Prevalence** | | | **Cumulative Incidence** | | |
| --- | --- | --- | --- | --- | --- | --- |
|  | **Intervention**  **Agents on PrEP** | **Intervention Agents not on PrEP** | **Control Agents** | **Intervention**  **Agents on PrEP** | **Intervention Agents not on PrEP** | **Control Agents** |
| **Among Components with *M* = 1** | | | | | | |
| Main | 5.5 | 357.1 | 404.2 | 5.5 | 19.0 | 67.9 |
| *PrEP Adherence* |  |  |  |  |  |  |
| 80% (White MSM)  50% (Black MSM) | 6.5 | 356.3 | 408.9 | 6.5 | 19.1 | 68.4 |
| **Among Components with *M* = 0** | | | | | | |
| Main | 4.9 | 355.8 | 394.8 | 4.9 | 16.4 | 57.3 |
| *PrEP Adherence* |  |  |  |  |  |  |
| 80% (White MSM)  50% (Black MSM) | 5.7 | 357.3 | 398.9 | 5.7 | 16.7 | 57.9 |

**Supplementary Table S10.** Sensitivity analyses of PrEP adherence, HIV prevalence and incidence in a two-stage randomization with 70% coverage in the intervention group in an agent-based model representing among men who have sex with men Atlanta, Georgia, 2015-2017, stratified by the effect modifier $M$ (density).

| **Scenario** | **HIV Prevalence** | | | **Cumulative Incidence** | | |
| --- | --- | --- | --- | --- | --- | --- |
|  | **Intervention**  **Agents on PrEP** | **Intervention Agents not on PrEP** | **Control Agents** | **Intervention**  **Agents on PrEP** | **Intervention Agents not on PrEP** | **Control Agents** |
| **Among Components with *M* = 1** | | | | | | |
| Main | 2.8 | 210.3 | 232.8 | 2.9 | 9.6 | 33.0 |
| *PrEP Adherence* |  |  |  |  |  |  |
| 80% (White MSM)  50% (Black MSM) | 3.4 | 211.2 | 235.1 | 3.4 | 9.7 | 33.4 |
| **Among Components with *M* = 0** | | | | | | |
| Main | 7.5 | 502.6 | 566.2 | 7.5 | 25.8 | 92.2 |
| *PrEP Adherence* |  |  |  |  |  |  |
| 80% (White MSM)  50% (Black MSM) | 8.8 | 502.4 | 572.7 | 8.8 | 26.1 | 92.9 |

**Supplementary Table S11.** Sensitivity analyses of PrEP discontinuation, HIV prevalence and incidence in a two-stage randomization with 70% coverage in the intervention group in an agent-based model representing among men who have sex with men Atlanta, Georgia, 2015-2017, stratified by the effect modifier $M$ (drug use).

| **Scenario** | **HIV Prevalence** | | | **Cumulative Incidence** | | |
| --- | --- | --- | --- | --- | --- | --- |
|  | **Intervention**  **Agents on PrEP** | **Intervention Agents not on PrEP** | **Control Agents** | **Intervention**  **Agents on PrEP** | **Intervention Agents not on PrEP** | **Control Agents** |
| **Among Components with *M* = 1** | | | | | | |
| Main | 4.0 | 291.8 | 327.4 | 4.0 | 14.3 | 51.0 |
| *PrEP Discontinuation* |  |  |  |  |  |  |
| 10% | 24.6 | 287.5 | 323.9 | 24.6 | 13.4 | 50.4 |
| **Among Components with *M* = 0** | | | | | | |
| Main | 6.4 | 421.1 | 471.6 | 6.4 | 21.1 | 74.2 |
| *PrEP Discontinuation* |  |  |  |  |  |  |
| 10% | 36.0 | 414.0 | 467.6 | 36.0 | 21.4 | 73.5 |

**Supplementary Table S12.** Sensitivity analyses of PrEP discontinuation, HIV prevalence and incidence in a two-stage randomization with 70% coverage in the intervention group in an agent-based model representing among men who have sex with men Atlanta, Georgia, 2015-2017, stratified by the effect modifier $M$ (HIV prevalence).

| **Scenario** | **HIV Prevalence** | | | **Cumulative Incidence** | | |
| --- | --- | --- | --- | --- | --- | --- |
|  | **Intervention**  **Agents on PrEP** | **Intervention Agents not on PrEP** | **Control Agents** | **Intervention**  **Agents on PrEP** | **Intervention Agents not on PrEP** | **Control Agents** |
| **Among Components with *M* = 1** | | | | | | |
| Main | 4.4 | 358.0 | 389.3 | 4.4 | 13.8 | 47.7 |
| *PrEP Discontinuation* |  |  |  |  |  |  |
| 10% | 23.5 | 351.1 | 385.4 | 23.5 | 13.8 | 47.2 |
| **Among Components with *M* = 0** | | | | | | |
| Main | 6.0 | 354.9 | 409.8 | 6.0 | 21.6 | 77.5 |
| *PrEP Discontinuation* |  |  |  |  |  |  |
| 10% | 37.1 | 350.4 | 406.1 | 37.1 | 22.1 | 76.8 |

**Supplementary Table S13.** Sensitivity analyses of PrEP discontinuation, HIV prevalence and incidence in a two-stage randomization with 70% coverage in the intervention group in an agent-based model representing among men who have sex with men Atlanta, Georgia, 2015-2017, stratified by the effect modifier $M$ (bridging potential).

| **Scenario** | **HIV Prevalence** | | | **Cumulative Incidence** | | |
| --- | --- | --- | --- | --- | --- | --- |
|  | **Intervention**  **Agents on PrEP** | **Intervention Agents not on PrEP** | **Control Agents** | **Intervention**  **Agents on PrEP** | **Intervention Agents not on PrEP** | **Control Agents** |
| **Among Components with *M* = 1** | | | | | | |
| Main | 5.5 | 357.1 | 404.2 | 5.5 | 19.0 | 67.9 |
| *PrEP Discontinuation* |  |  |  |  |  |  |
| 10% | 32.7 | 350.6 | 400.1 | 32.7 | 19.3 | 67.3 |
| **Among Components with *M* = 0** | | | | | | |
| Main | 4.9 | 355.8 | 394.8 | 4.9 | 16.4 | 57.3 |
| *PrEP Discontinuation* |  |  |  |  |  |  |
| 10% | 27.9 | 350.8 | 391.4 | 27.9 | 16.6 | 56.7 |

**Supplementary Table S14.** Sensitivity analyses of PrEP discontinuation, HIV prevalence and incidence in a two-stage randomization with 70% coverage in the intervention group in an agent-based model representing among men who have sex with men Atlanta, Georgia, 2015-2017, stratified by the effect modifier $M$ (density).

| **Scenario** | **HIV Prevalence** | | | **Cumulative Incidence** | | |
| --- | --- | --- | --- | --- | --- | --- |
|  | **Intervention**  **Agents on PrEP** | **Intervention Agents not on PrEP** | **Control Agents** | **Intervention**  **Agents on PrEP** | **Intervention Agents not on PrEP** | **Control Agents** |
| **Among Components with *M* = 1** | | | | | | |
| Main | 2.8 | 210.3 | 232.8 | 2.9 | 9.6 | 33.0 |
| *PrEP Discontinuation* |  |  |  |  |  |  |
| 10% | 16.1 | 208.1 | 230.0 | 16.1 | 9.7 | 32.5 |
| **Among Components with *M* = 0** | | | | | | |
| Main | 7.5 | 502.6 | 566.2 | 7.5 | 25.8 | 92.2 |
| *PrEP Discontinuation* |  |  |  |  |  |  |
| 10% | 44.5 | 493.4 | 561.5 | 44.5 | 26.2 | 91.5 |

**Supplementary Table S15.** Sensitivity analyses with lower PrEP adherence or PrEP discontinuation, estimated spillover risk differences (RD) and risk ratios (RR) with 95% simulation intervals (SI) for HIV cumulative incidence in a two-stage randomization with 70% coverage in the intervention group in an agent-based model representing among men who have sex with men Atlanta, Georgia, 2015-2017, stratified by the effect modifier $M$ (drug use).

| **Scenario** | RD (95% SI) | RR (95% SI) |
| --- | --- | --- |
| **Among Components with *M* = 1** | | |
| Main | -0.07 (-0.14, 0.00) | 0.53 (0.20, 1.0) |
| *PrEP Adherence* |  |  |
| 80% (White MSM)  50% (Black MSM) | -0.07 (-0.13, 0.00) | 0.53 (0.21, 1.0) |
| *PrEP Discontinuation* |  |  |
| 10% | -0.07 (-0.14, -0.001) | 0.51 (0.18, 0.99) |
| **Among Components with *M* = 0** | | |
| Main | -0.08 (-0.14, -0.13) | 0.47 (0.24, 0.78) |
| *PrEP Adherence* |  |  |
| 80% (White MSM)  50% (Black MSM) | -0.08 (-0.14, -0.03) | 0.48 (0.24, 0.80) |
| *PrEP Discontinuation* |  |  |
| 10% | -0.09 (-0.14, -0.03) | 0.46 (0.22, 0.79) |

**Supplementary Table S16.** Sensitivity analyses with lower PrEP adherence or PrEP discontinuation, estimated spillover risk differences (RD) and risk ratios (RR) with 95% simulation intervals (SI) for HIV cumulative incidence in a two-stage randomization with 70% coverage in the intervention group in an agent-based model representing among men who have sex with men Atlanta, Georgia, 2015-2017, stratified by the effect modifier $M$ (HIV prevalence).

| **Scenario** | RD (95% SI) | RR (95% SI) |
| --- | --- | --- |
| **Among Components with *M* = 1** | | |
| Main | -0.13 (-0.22, -0.04) | 0.41 (0.18, 0.76) |
| *PrEP Adherence* |  |  |
| 80% (White MSM)  50% (Black MSM) | -0.13 (-0.22, -0.04) | 0.43 (0.18, 0.76) |
| *PrEP Discontinuation* |  |  |
| 10% | -0.13 (-0.22, -0.03) | 0.41 (0.16, 0.81) |
| **Among Components with *M* = 0** | | |
| Main | -0.05 (-0.10, -0.01) | 0.56 (0.26, 0.91) |
| *PrEP Adherence* |  |  |
| 80% (White MSM)  50% (Black MSM) | -0.05 (-0.10, 0.00) | 0.56 (0.25, 0.98) |
| *PrEP Discontinuation* |  |  |
| 10% | -0.05 (-0.10, -0.004) | 0.54 (0.26, 0.96) |

**Supplementary Table S17.** Sensitivity analyses with lower PrEP adherence or PrEP discontinuation, estimated spillover risk differences (RD) and risk ratios (RR) with 95% simulation intervals (SI) for HIV cumulative incidence in a two-stage randomization with 70% coverage in the intervention group in an agent-based model representing among men who have sex with men Atlanta, Georgia, 2015-2017, stratified by the effect modifier $M$ (bridging potential).

| **Scenario** | RD (95% SI) | RR (95% SI) |
| --- | --- | --- |
| **Among Components with *M* = 1** | | |
| Main | -0.04 (-0.09, 0.02) | 0.68 (0.31, 1.2) |
| *PrEP Adherence* |  |  |
| 80% (White MSM)  50% (Black MSM) | -0.04 (-0.10, 0.02) | 0.68 (0.31, 1.2) |
| *PrEP Discontinuation* |  |  |
| 10% | -0.05 (-0.11, 0.02) | 0.63 (0.26, 1.16) |
| **Among Components with *M* = 0** | | |
| Main | -0.08 (-0.12, -0.03) | 0.44 (0.23, 0.72) |
| *PrEP Adherence* |  |  |
| 80% (White MSM)  50% (Black MSM) | -0.07 (-0.12, -0.03) | 0.44 (0.23, 0.73) |
| *PrEP Discontinuation* |  |  |
| 10% | -0.08 (-0.12, -0.03) | 0.44 (0.21, 0.75) |

**Supplementary Table S18.** Sensitivity analyses with lower PrEP adherence or PrEP discontinuation, estimated spillover risk differences (RD) and risk ratios (RR) with 95% simulation intervals (SI) for HIV cumulative incidence in a two-stage randomization with 70% coverage in the intervention group in an agent-based model representing among men who have sex with men Atlanta, Georgia, 2015-2017, stratified by the effect modifier $M$ (density).

| **Scenario** | RD (95% SI) | RR (95% SI) |
| --- | --- | --- |
| **Among Components with *M* = 1** | | |
| Main | -0.08 (-0.14, -0.02) | 0.42 (0.17, 0.79) |
| *PrEP Adherence* |  |  |
| 80% (White MSM)  50% (Black MSM) | -0.08 (-0.13, -0.02) | 0.42 (0.16, 0.79) |
| *PrEP Discontinuation* |  |  |
| 10% | -0.08 (-0.14, -0.02) | 0.42 (0.15, 0.80) |
| **Among Components with *M* = 0** | | |
| Main | -0.06 (-0.10, -0.01) | 0.58 (0.30, 0.94) |
| *PrEP Adherence* |  |  |
| 80% (White MSM)  50% (Black MSM) | -0.06 (-0.10, -0.01) | 0.59 (0.32, 0.95) |
| *PrEP Discontinuation* |  |  |
| 10% | -0.06 (-0.11, -0.01) | 0.55 (0.29, 0.93) |

**Supplementary Figure S1.** Estimated risk difference spillover effects of PrEP on cumulative incidence of HIV by four modifiers (M = 1 if prevalence above median vs. M = 0 at or below median) in two-stage randomized designs of a pre-exposure prophylaxis (PrEP) intervention with 70% coverage in an agent-based model representing men who have sex with men, Atlanta, Georgia, 2015-2017 and PrEP adherence set to 50% among Black MSM and 80% among White MSM**.** Lines within boxes, median values; box borders, interquartile ranges (75th and 25th percentiles); bars, 90th and 10th percentiles; points, outliers. Shaded shape represented the distribution of estimates and dashed lines represent the null value. (n = 3,953).

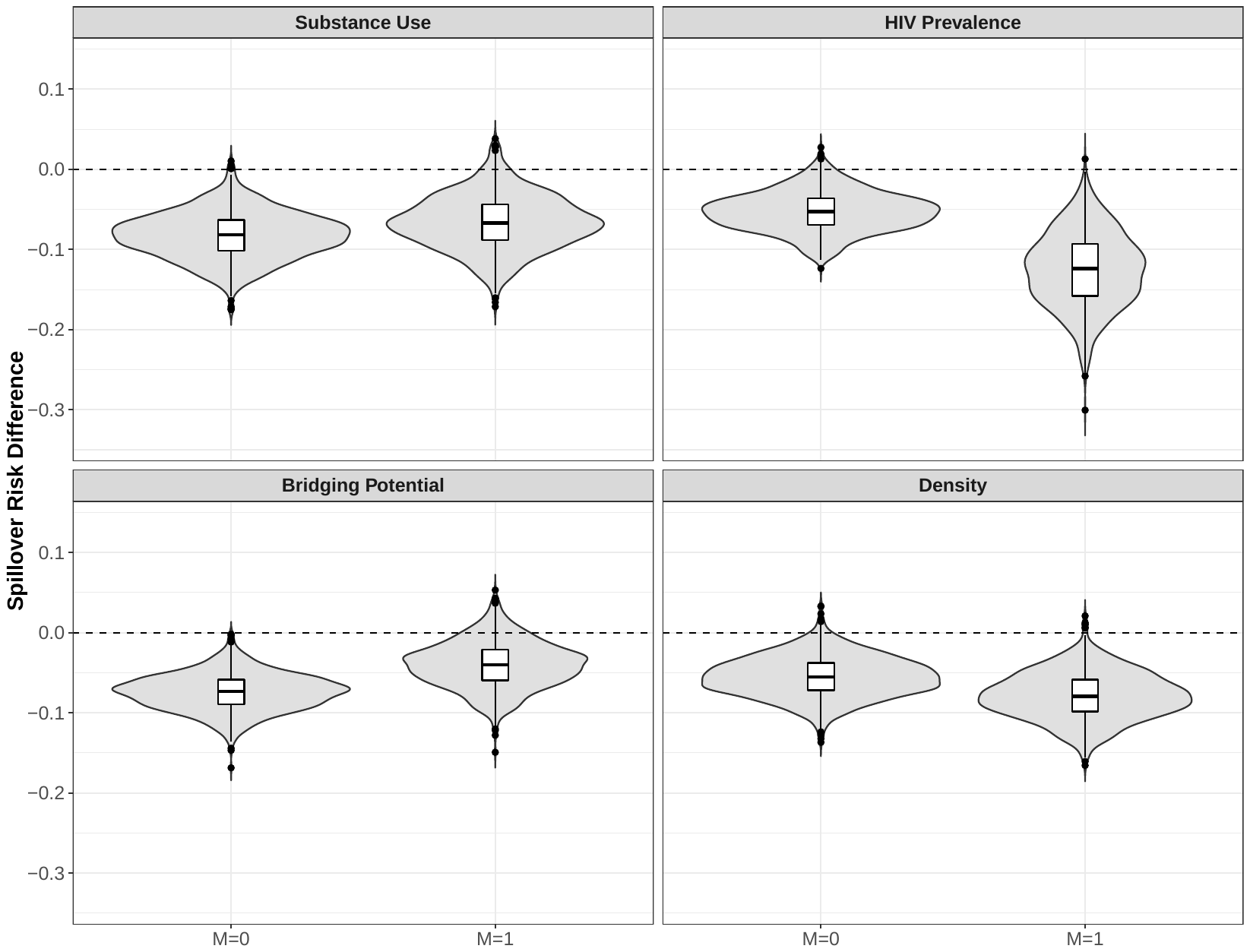

**Supplementary Figure S2.** Estimated risk ratio spillover effects of PrEP on cumulative incidence of HIV by four modifiers (M = 1 if prevalence above median vs. M = 0 at or below median) in two-stage randomized designs of a pre-exposure prophylaxis (PrEP) intervention with 70% coverage in an agent-based model representing men who have sex with men, Atlanta, Georgia, 2015-2017 and PrEP adherence set to 50% among Black MSM and 80% among White MSM. Lines within boxes, median values; box borders, interquartile ranges (75th and 25th percentiles); bars, 90th and 10th percentiles; points, outliers. Shaded shape represented the distribution of estimates and dashed lines represent the null value. (n = 3,953).

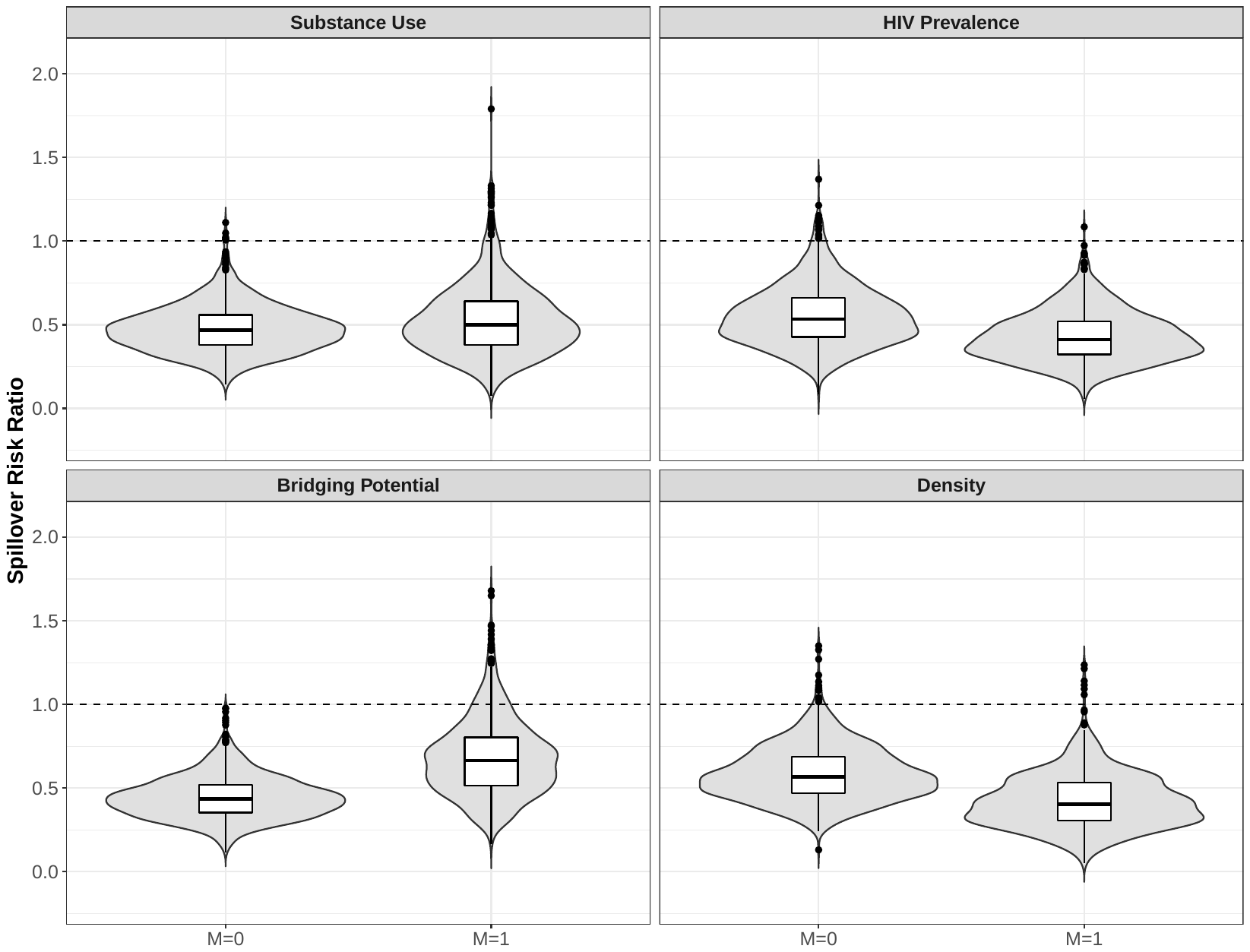

**Supplementary Figure S3.** Estimated risk difference spillover effects of PrEP on cumulative incidence of HIV by four modifiers (M = 1 if prevalence above median vs. M = 0 at or below median) in two-stage randomized designs of a pre-exposure prophylaxis (PrEP) intervention with 70% coverage in an agent-based model representing men who have sex with men, Atlanta, Georgia, 2015-2017 and PrEP discontinuation in each monthly interval set to 10%**.** Lines within boxes, median values; box borders, interquartile ranges (75th and 25th percentiles); bars, 90th and 10th percentiles; points, outliers. Shaded shape represented the distribution of estimates and dashed lines represent the null value. (n = 3,896).

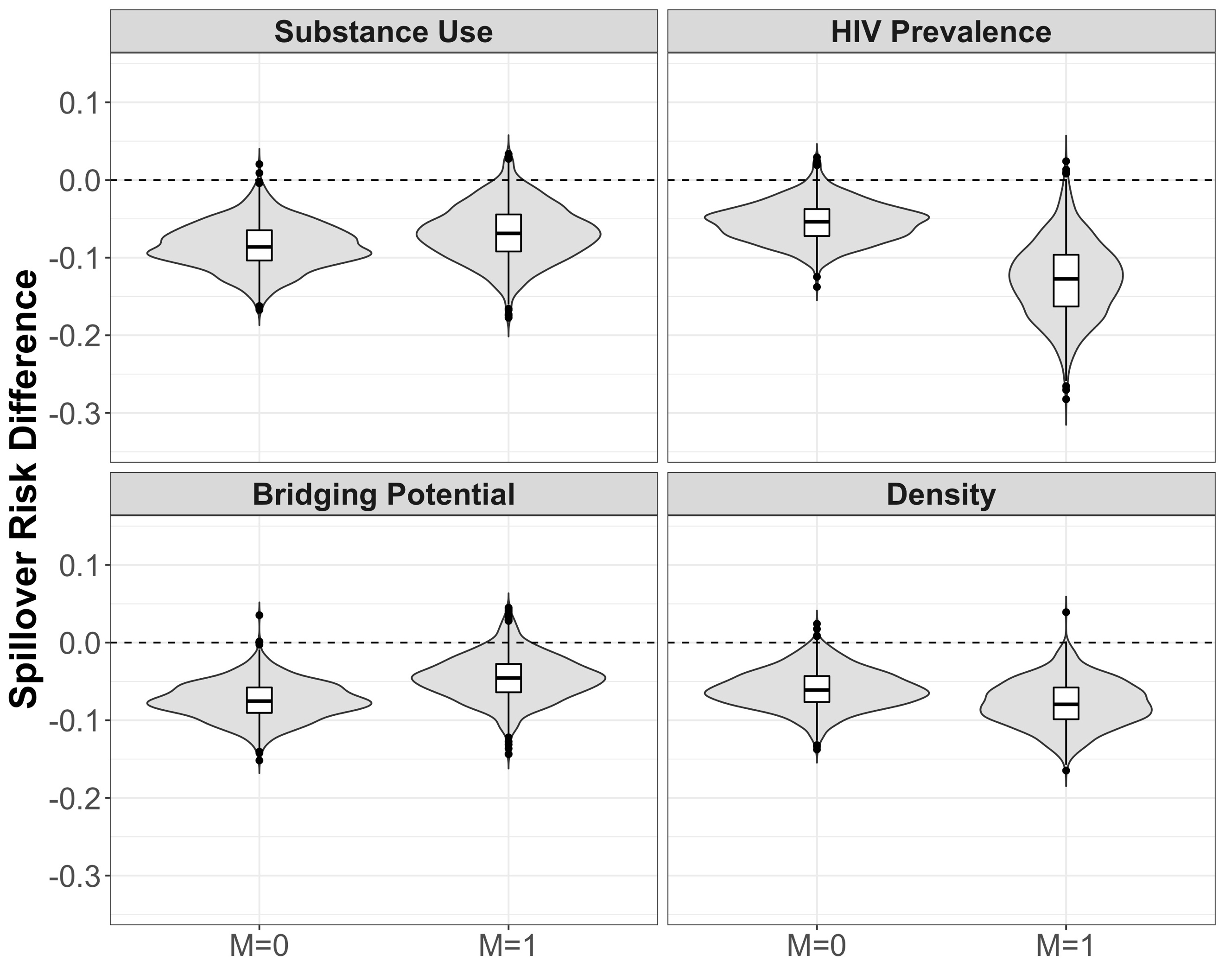

**Supplementary Figure S4.** Estimated risk ratio spillover effects of PrEP on cumulative incidence of HIV by four modifiers (M = 1 if prevalence above median vs. M = 0 at or below median) in two-stage randomized designs of a pre-exposure prophylaxis (PrEP) intervention with 70% coverage in an agent-based model representing men who have sex with men, Atlanta, Georgia, 2015-2017 and PrEP discontinuation in each monthly interval set to 10%. Lines within boxes, median values; box borders, interquartile ranges (75th and 25th percentiles); bars, 90th and 10th percentiles; points, outliers. Shaded shape represented the distribution of estimates and dashed lines represent the null value. (n = 3,896).

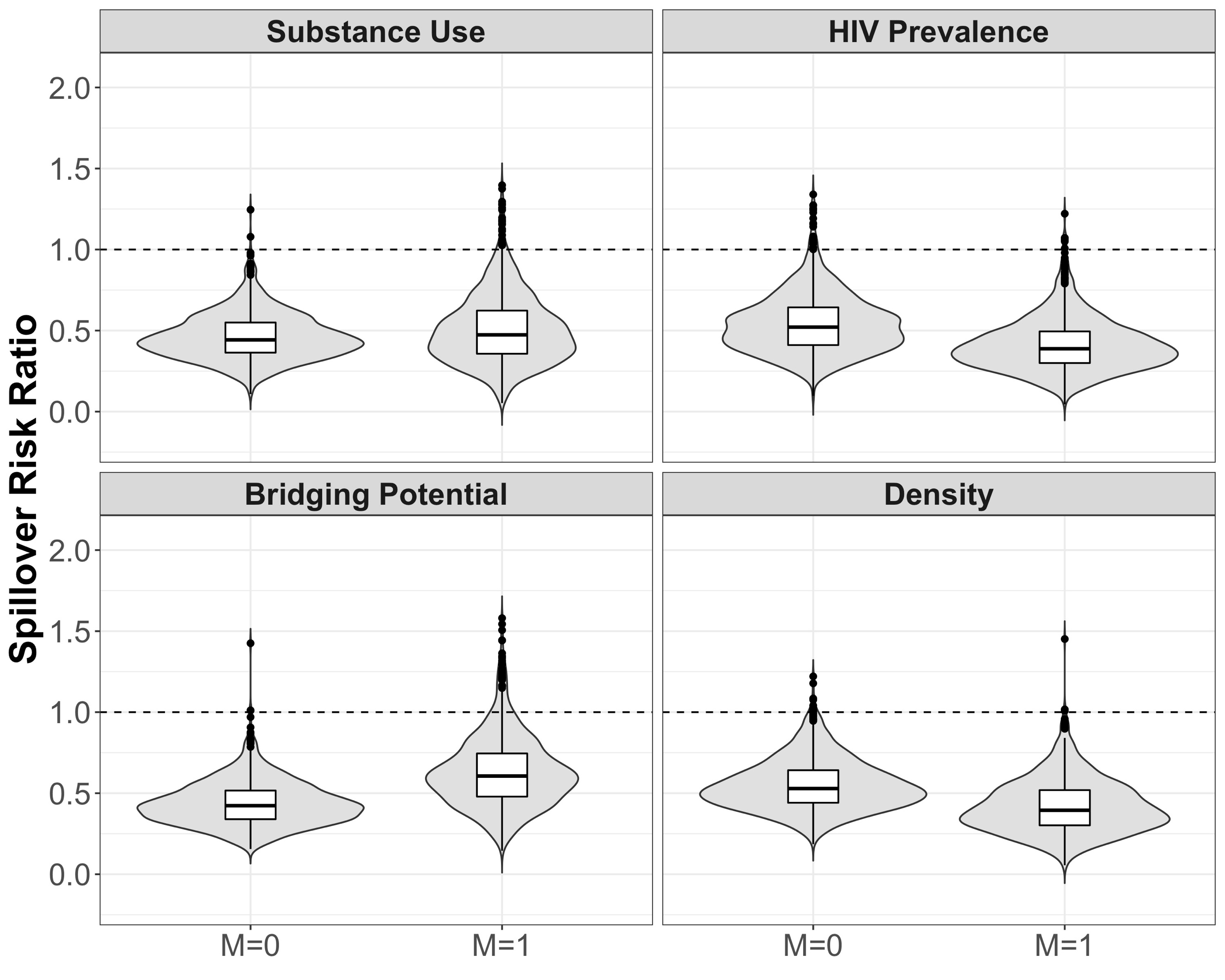

39.	Van Rossum G, Drake FL Jr. *Python Reference Manual*. Amsterdam, the Netherlands: Center for Mathematics and Computer Science; 1995.28.

40.	Walt Svd, Colbert SC, Varoquaux G. The NumPy array: a structure for efficient numerical computation. *Computing in Science & Engineering*. 2011;13(2):22-30.

41.	Hagberg AA, Schult DA, Swart PJ. Exploring network structure, dynamics, and function using NetworkX. In: Varoquaux G, Vaught T, Millman J, eds. Proceedings of the 7th Python in Science Conference (SciPy 2008)—Austin, Texas, June 28–July 3, 2008. Austin, TX: SciPy; 2008:11–15. http://conference.scipy.org/proceedings/scipy2008/. Accessed May 28, 2019.30.

42.	R Core Team. *R: A Language and Environment for Statistical Computing*. Vienna, Austria: R Foundation for Statistical Computing; 2014. http://www.R-project.org. Accessed July 9, 2019.35.

43.	Wickham H. ggplot2: Elegant Graphics for Data Analysis. New York, NY: Springer Publishing Company; 2009.
